# COVID-19-associated AKI in hospitalized US patients: incidence, temporal trends, geographical distribution, risk factors and mortality

**DOI:** 10.1101/2022.09.02.22279398

**Authors:** Masthead, Yun Jae Yoo, Kenneth J. Wilkins, Fadhl Alakwaa, Feifan Liu, Luke A. Torre-Healy, Spencer Krichevsky, Stephanie S. Hong, Ankit Sakhuja, Chetan K. Potu, Joel H. Saltz, Rajiv Saran, Richard L. Zhu, Soko Setoguchi, Sandra L. Kane-Gill, Sandeep K. Mallipattu, Yongqun He, David H. Ellison, James Brian Byrd, Chirag R. Parikh, Richard A. Moffitt, Farrukh M. Koraishy

## Abstract

**Background:** Acute kidney injury (AKI) is associated with mortality in patients hospitalized with COVID-19, however, its incidence, geographic distribution, and temporal trends since the start of the pandemic are understudied.

**Methods:** Electronic health record data were obtained from 53 health systems in the United States (US) in the National COVID Cohort Collaborative (N3C). We selected hospitalized adults diagnosed with COVID-19 between March 6th, 2020, and January 6th, 2022. AKI was determined with serum creatinine (SCr) and diagnosis codes. Time were divided into 16-weeks (P1-6) periods and geographical regions into Northeast, Midwest, South, and West. Multivariable models were used to analyze the risk factors for AKI or mortality.

**Results:** Out of a total cohort of 306,061, 126,478 (41.0 %) patients had AKI. Among these, 17.9% lacked a diagnosis code but had AKI based on the change in SCr. Similar to patients coded for AKI, these patients had higher mortality compared to those without AKI. The incidence of AKI was highest in P1 (49.3%), reduced in P2 (40.6%), and relatively stable thereafter. Compared to the Midwest, the Northeast, South, and West had higher adjusted AKI incidence in P1, subsequently, the South and West regions continued to have the highest relative incidence. In multivariable models, AKI defined by either SCr or diagnostic code, and the severity of AKI was associated with mortality.

**Conclusions:** Uncoded cases of COVID-19-associated AKI are common and associated with mortality. The incidence and distribution of COVID-19-associated AKI have changed since the first wave of the pandemic in the US.

## Introduction

Coronavirus Disease 2019 (COVID-19) caused by the novel Severe Acute Respiratory Syndrome Coronavirus-2 (SARS-CoV-2) is the most significant global pandemic of the 21^st^ century, with a devastating impact on individuals and society worldwide^1^. Patients hospitalized with COVID-19 can develop multi-organ dysfunction including acute kidney injury (AKI). AKI is a common comorbid condition and a major predictor of death in patients hospitalized with COVID-19^2-5^. The severity of AKI has been associated with in-hospital complications and mortality^6,7^.

In electronic health record (EHR)-based datasets, where urine output measurements are not reliable, serum creatinine (SCr) and diagnostic codes are used to define AKI. Previous studies have shown the limitations in the use of EHR billing codes to capture the true incidence of AKI^8,9^. In a study of 10,056 patients hospitalized between 1996 and 2008, Grams et al. reported low sensitivity of billing codes to identify milder cases of AKI as compared with the 2012 Kidney Disease Improving Global Outcomes (KDIGO) serum creatinine (SCr)-based criteria^8^ and noted that code-based AKI captured more severe AKI cases with higher short-term mortality compared to SCr-based AKI cases. This suggests that mild AKI (stage 1) is often not captured with a billing code although it has been associated with adverse outcomes ^10,11^. On the other hand, the challenge in using the rise in SCr as an indicator of AKI lies in the lack of a baseline SCr for a significant number of hospitalized patients in an EHR dataset. Clinicians often have other sources of obtaining baseline SCr (not available in EHR data extraction) and might use the change in urine output to define AKI. Hence, a combination of using both diagnostic billing codes and change in SCr in EHR datasets might lead to a more accurate estimation of the incidence of AKI. The relative incidence of AKI that is documented via billing codes *versus* SCr-based criteria during hospitalizations with COVID-19, and the relative association of coded *versus* uncoded AKI cases with mortality, is unknown.

Since the start of the pandemic in December 2019, COVID-19 has shown wide variations in incidence, hospitalization and death rates worldwide^12,13^. Even within the United States (US) the incidence, severity, and mortality associated with COVID-19 have evolved significantly since March 2020^14,15^. In an early study of 5,216 US veterans (94% male) hospitalized with COVID-19, a decline in the incidence rate of AKI from 40% in March 2020 to 27% in July 2020 was reported along with a wide geographic variation of 10-56%^16^. Yet, nearly all studies of COVID-19 associated AKI to date have been relatively limited in sample sizes, geographic diversity, and observation periods. To address these limitations, we used the National COVID Cohort Collaborative (N3C^17^), currently the largest public-access enclave of patients with COVID-19 across the USA (nearly 5 million at the time of this study); with capacity to characterize COVID-19-associated AKI and its geographic heterogeneity period-by-period over the course of the pandemic.

We hypothesized that among patients hospitalized with COVID-19, both diagnosis codes and changes in SCr will be required to capture the incidence of AKI and that AKI incidence and mortality will vary with time and region.

## Methods

### N3C data: Ingestion & Harmonization

N3C brings together de-identified EHR data (dating back to January 1, 2018) from 72 healthcare institutions across the US into a centralized repository, allowing detailed study of a geographically diverse population of patients with COVID-19 from each participating site as well as matched controls^17^.

Institutions contributed EHR data to the N3C consortium using various source data models^18^, i.e., PCORnet, PEDSnet, ACT, TriNetX, and OMOP. All data were harmonized, and mapped to the OMOP Common Data Model (V5.3.1)^19^ in accordance with data quality and harmonization checks^20^. Characteristics of the adult and pediatric cohorts have previously been described^15,21^.

### Study Oversight

N3C has been approved by the National Institutes of Health’s Institutional Review Board (IRB). Each N3C contributor site maintains a data transfer agreement approved by its IRB. The analyses reported in this study were separately approved by the IRB of each participating institution. The IRB reviews included a waiver of informed consent.

### Definition of COVID-19 and other variables

COVID-19 was defined by a positive PCR or antigen test or by a COVID-19 diagnostic code within 7 days of hospitalization (**Figure 1**). **Supplementary Table 1** contains a list of all OMOP concepts used to define all variables used in our study and other variables are described in the Supplementary Methods.

**Figure 1.**
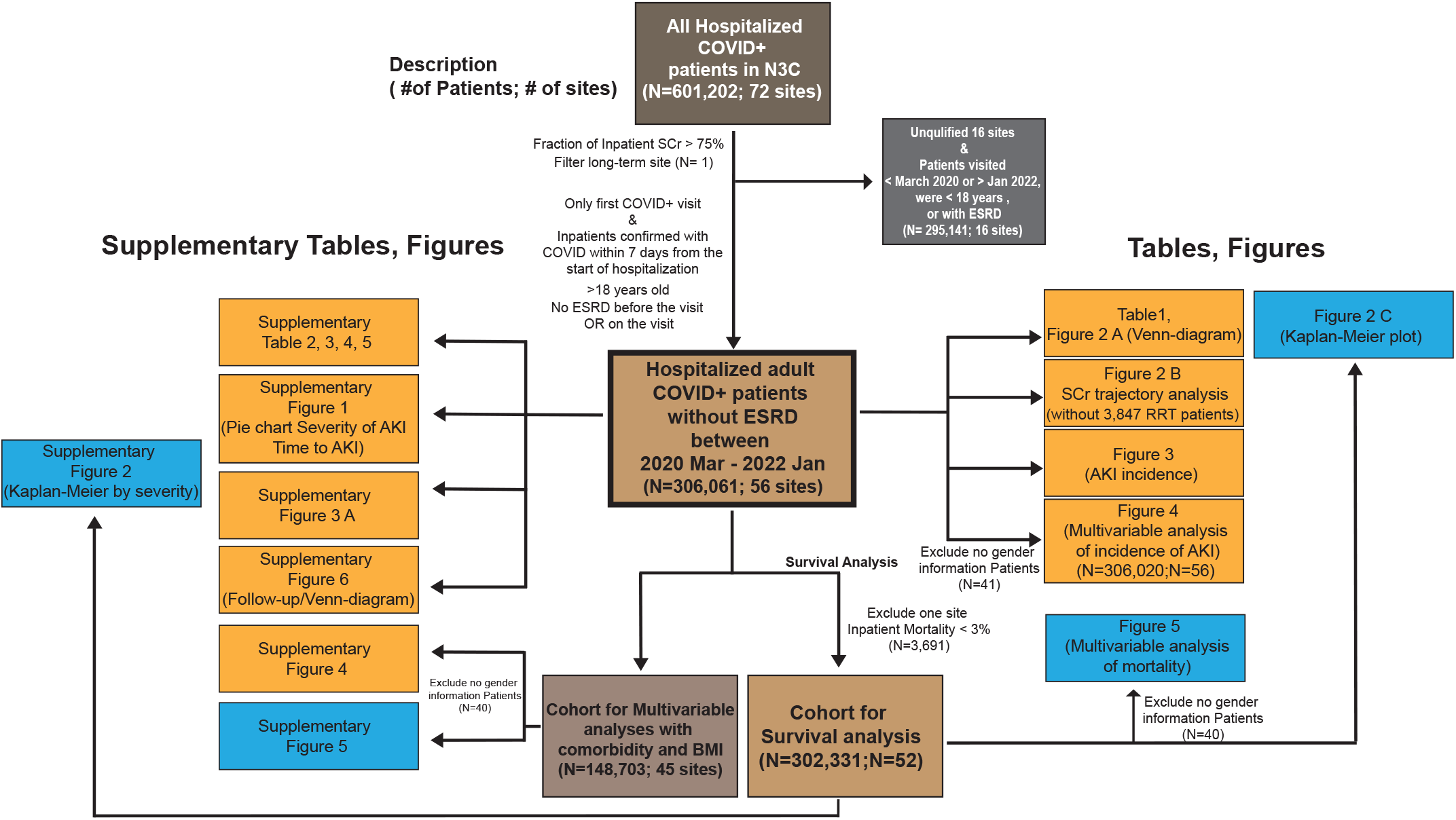
Flow diagram showing the total number of patients used in the study and the patients used in each table and figure. The study included all adult patients in the N3C data set with COVID-19 positive within 7 days of an inpatient hospitalization. Other patients were excluded for study due to site-specific data quality issues, pre-existing ESRD, or lack of SCr data.

### Cohort Definition, Inclusion and Exclusion Criteria

This study is a retrospective analysis of patients who: (1) were hospitalized between March 6^th^ 2020 through January 6^th^ 2022, (2) were at least 18 years old, (3) had a contemporary COVID-19 diagnosis, and (4) did not have a diagnosis of End-Stage Renal Disease (ESRD) on or before their COVID-19 associated hospitalization **(Figure 1)**. For all endpoints, March 6th, 2022 was the last day of follow-up. We built the analytic cohort from the April 2^nd^, 2022 release of N3C data, but additionally applied site-level data quality criteria as described in the Supplementary Methods.

After applying these inclusion and exclusion criteria for data partners, we further excluded patients with fewer than 2 SCr values, i.e., those unable to be assessed for AKI, unless they had a diagnostic code of AKI on their COVID-19 associated hospitalization (N= 78,526). Considering an initial population of 601,202 confirmed COVID-19 hospitalized patients from 72 sites, using the aforementioned criteria, our final cohort consisted of 306,061 adults with COVID-19 associated hospitalizations from 53 sites **(Figure 1)**.

### Renal Measures

#### Definition of AKI

We excluded 20,289 patients diagnosed with ESRD before or during the index hospitalization (Figure 1). Patients were required to have ≥2 SCr measurements available to diagnose AKI, except for 2,141 patients that carried a diagnosis code for AKI (Figure 2). Patients with any of the 25 diagnosis codes related to AKI (Supplementary Table 1) were designated as “**Code-based AKI**”.

**Figure 2.**
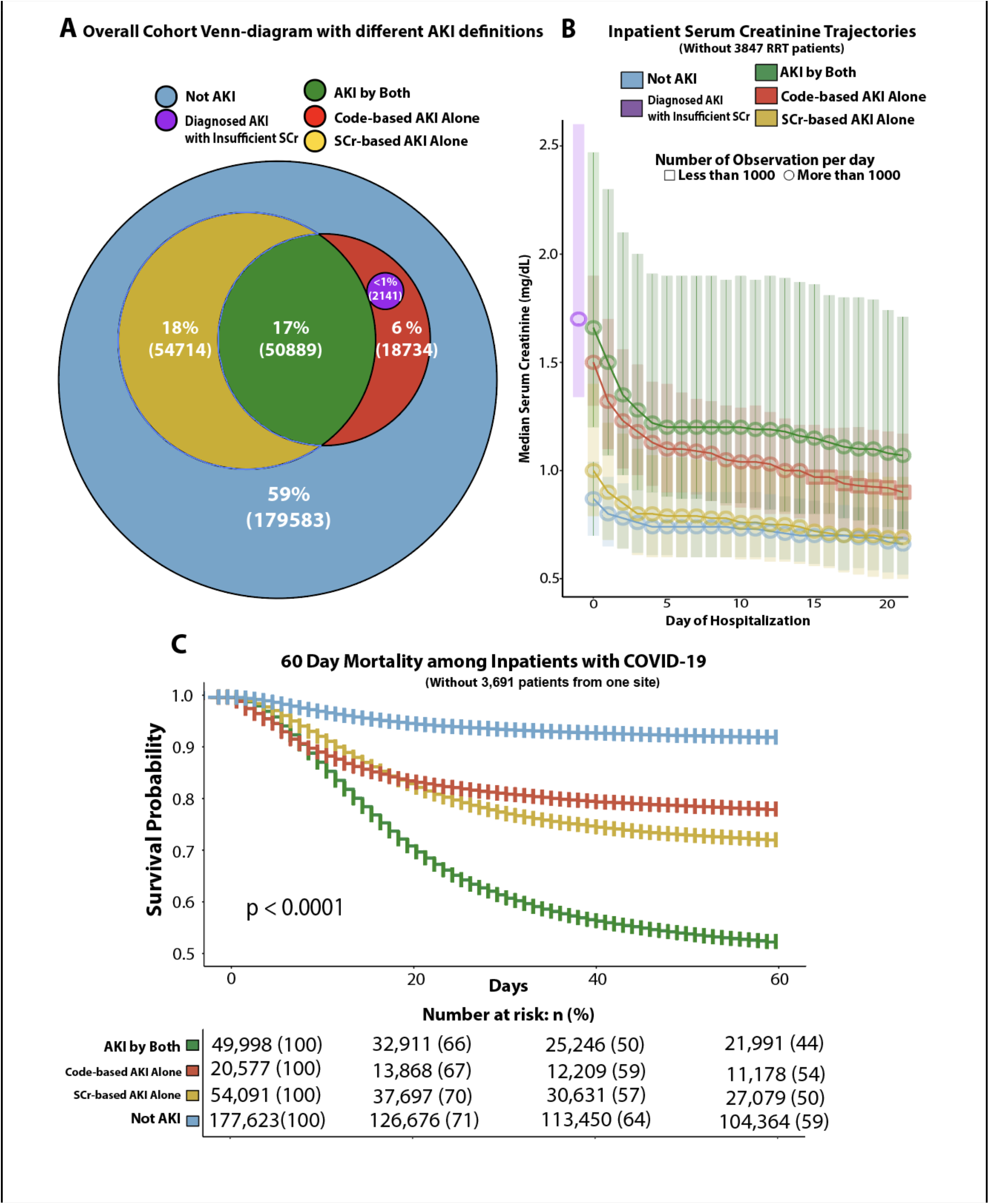
Development of the cohort, definitions of COVID-19-associated AKI and comparative mortality rates. (a) Overall prevalence of AKI by different AKI definitions in a total of 306,061 adult cohorts. (b) SCr trajectories (excluding patient who underwent acute dialysis) by day of admission between patient groups classified in different AKI definitions. (c) 60-day survival after diagnosis of COVID-19 in the inpatient population, shown as stratum-specific Kaplan-Meier curves in a color scheme matching (a) and (b), with pvalue for the log-rank test of differences among strata).

Three definitions were used to define “**SCr-based AKI**”.

1. When the difference in SCr value increased by more than 0.3 mg/dL within any 48 hour period during hospitalization.
2. When the SCr value increased by more than 1.5 times from baseline within any 7 day period during hospitalization.
3. When the patient’s maximum SCr value during hospitalization was higher than their baseline SCr value (defined in Supplementary Methods) by at least 1.5 times.

If any one of the above three definitions was satisfied, we classified it as “**SCr-based AKI”**. The number of patients for each SCr-based AKI definition is shown in **Supplementary Figure 1A**. Unless otherwise noted, ‘**AKI**’ is this study was defined as meeting the criteria for either “SCr-based AKI” or “code-based AKI”,

Any patient requiring renal replacement therapy (RRT) was classified at Stage 3 AKI. The severity of AKI is further explained in the Supplementary Methods.

### Geographical Regions and Time Periods

Patients included in the study had their index hospitalization with COVID-19 between March 6^th^ 2020 and January 6^th^ 2022. The cohort, representing 96 weeks of enrollment, was divided into six equal 16-week intervals (sextiles): P1: March 6, 2020 - June 25, 2020, P2: - October 16, 2020, P3: - February 5, 2021, P4: - May 28, 2021, P5: - September 16, 2022, P6: -January 6, 2022. The cohort was also divided based on the location of the health-care sites into four geographic regions of the US (**Figure 3B**): West (10 sites, 26,522 patients), Northeast (12 sites, 56,092 patients), Midwest (18 sites, 166,456 patients), and South (12 sites, 56,871 patients).

**Figure 3.**
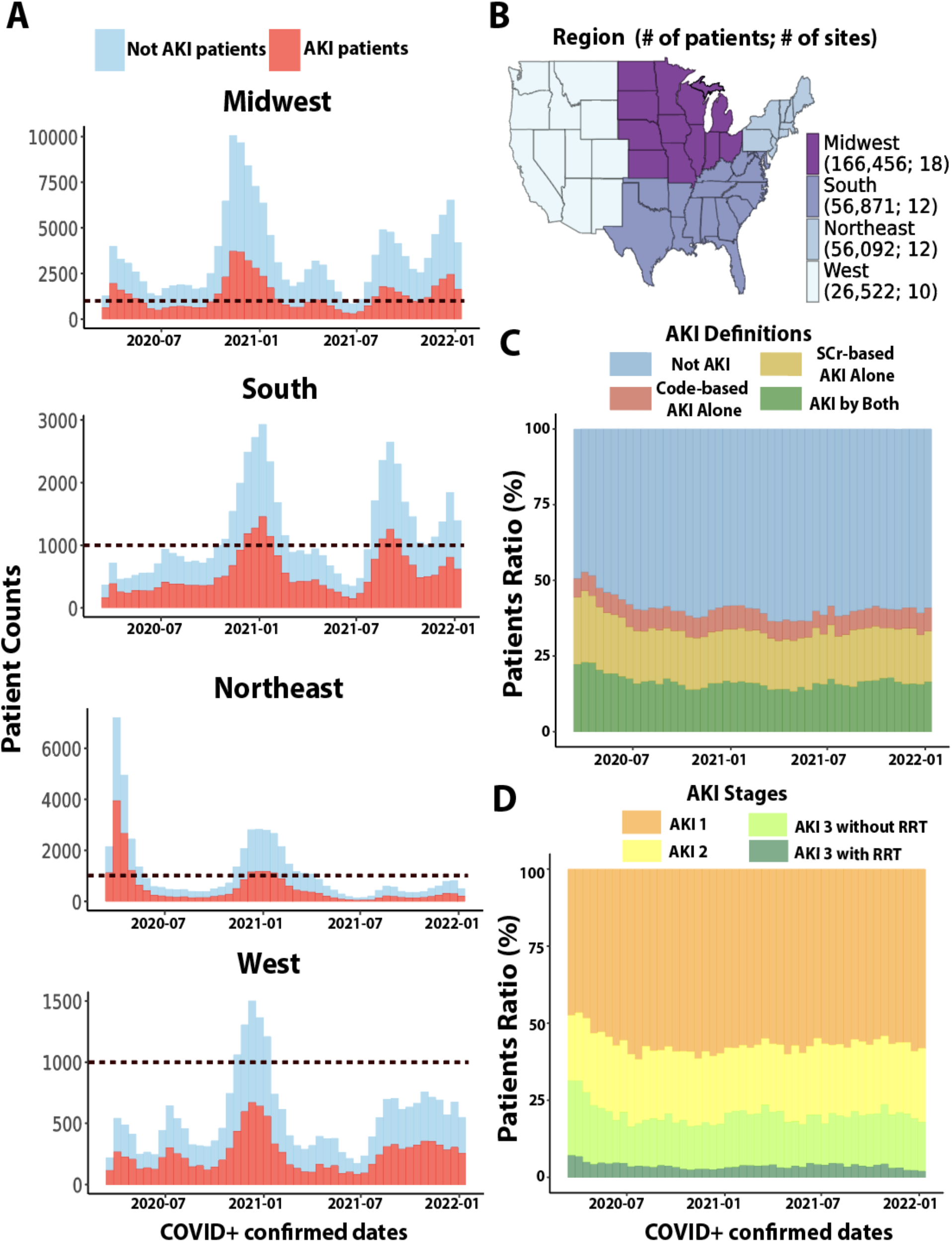
Temporal and geographical distribution of COVID-19-associated AKI.

### Statistical Analysis

The cohort construction for primary and subgroup analyses is described in **Figure 1**. Comparison of categorical variables was performed using Chi-square tests and for continuous variables, Student’s *t*-tests or ANOVA were used (**Table 1, Supplementary Table 2, 3, 4, 5**). All survival analyses were performed using the COVID-19 index date as a start date (earliest diagnosis date or positive test date), and death date as an endpoint (**Figure 2C & 5, Supplementary Figure 2**). A total of 306,061 patients (primary cohort) were used to define the geographic distribution and temporal trends of AKI (**Figure 3**). Multivariable logistic regression estimated the risk of AKI, quantifying relative risk in terms of odds ratios (ORs), and the Cox Proportional Hazard (CoxPH) model estimated mortality risk in terms of the hazard ratio (HR), with AKI-specific HRs for mortality as the primary endpoint. (**Figure 4 & 5, Supplementary Figure 4 & 5**). All analysis and visualization were done in the N3C enclave using SQL, Python, and R, including ggplot2^22^, survival^23^, and survminer^24^ packages. Further statistical details are provided in the Supplementary Methods.

**Table 1.** Descriptive characteristics of hospitalized COVID-19 positive patients with and without AKI.

| **Variables** | **Total**<br>**(N=306,061)** | **AKI**<br>**[Code-based OR**<br>**SCr-based]**<br>**(N= 126,478)** | **Not AKI**<br>**(N= 179,583)** | **p values** |
| --- | --- | --- | --- | --- |
| **Demographics** |  |  |  |  |
| **Age, Mean (SD)** | 61.71 (18.83) | 65.13 (16.59) | 59.3 (19.91) | < 0.0001 |
| **Gender (N, %)** |  |  |  |  |
| Female | 146,647 (47.91) | 55,319 (43.74) | 91,328 (50.86) |  |
| Male | 159,373 (52.07) | 71,141 (56.25) | 88,232 (49.13) | < 0.0001 |
| **Race (N, %)** |  |  |  |  |
| White | 192,482 (62.89) | 75,705 (59.86) | 116,777 (65.03) |  |
| Black | 58,564 (19.13) | 28,292 (22.37) | 30,272 (16.86) |  |
| Asian | 7,170 (2.34) | 3,067 (2.42) | 4,103 (2.28) |  |
| Others | 7,014 (2.29) | 2,736 (2.16) | 4,278 (2.38) | < 0.0001 |
| No Information | 40,831 (13.34) | 16,678 (13.19) | 24,153 (13.45) |  |
| **Ethnicity (N, %)** |  |  |  |  |
| Not Hispanic or Latino | 239,230 (78.16) | 99,666 (78.8) | 139,564 (77.72) |  |
| Hispanic or Latino | 41,220 (13.47) | 15,754 (12.46) | 25,466 (14.18) | < 0.0001 |
| No Information | 25,611 (8.37) | 11,058 (8.74) | 14,553 (8.1) |  |
| **Co-morbid conditions**<br>**(N, %)** |  |  |  |  |
| History available | 232,329 (75.91) | 92,551 (73.18) | 139,778 (77.83) | < 0.0001 |
| Among those with<br>histories available |  |  |  |  |
| CVD | 91,930 (39.57) | 43,291 (46.78) | 48,639 (34.8) | < 0.0001 |
| DM | 70,494 (30.34) | 34,731 (37.53) | 35,763 (25.59) | < 0.0001 |
| HF | 37,677 (16.22) | 20,019 (21.63) | 17,658 (12.63) | < 0.0001 |
| HTN | 120,353 (51.8) | 55,598 (60.07) | 64,755 (46.33) | < 0.0001 |
| **BMI, Mean (SD)** | 30.83 (8.62) | 30.65 (8.66) | 30.97 (8.58) | = 1.0000 |
| **Severity of illness (N, %)** |  |  |  |  |
| Sepsis | 58,506 (19.12) | 39,360 (31.12) | 19,146 (10.66) | < 0.0001 |
| IMV | 32,457 (10.6) | 28,511 (22.54) | 3,946 (2.2) | < 0.0001 |
| Length of hospital stay, Mean, days (IQRs) | 9.73 (7.0) | 14.39 (13.0) | 6.46 (5.0) |  |
| **Medications (N, %)** |  |  |  |  |
| VP | 39,907 (13.04) | 30,163 (23.85) | 9,744 (5.43) | < 0.0001 |
| **Death (N, %)** | 46,122 (15.07) | 33,735 (26.67) | 12,387 (6.9) | < 0.0001 |

**Abbreviations:**
DM (Diabetes Mellitus), HF (Heart Failure), HTN (Hypertension), CVD (Cardiovascular disease), BMI (Body Mass Index, kg/m<sup>2</sup>), IMV (Invasive Mechanical Ventilation), VP (vasopressors) IQR (Inter-quartile range), SD (standard deviation)

**Figure 4.**
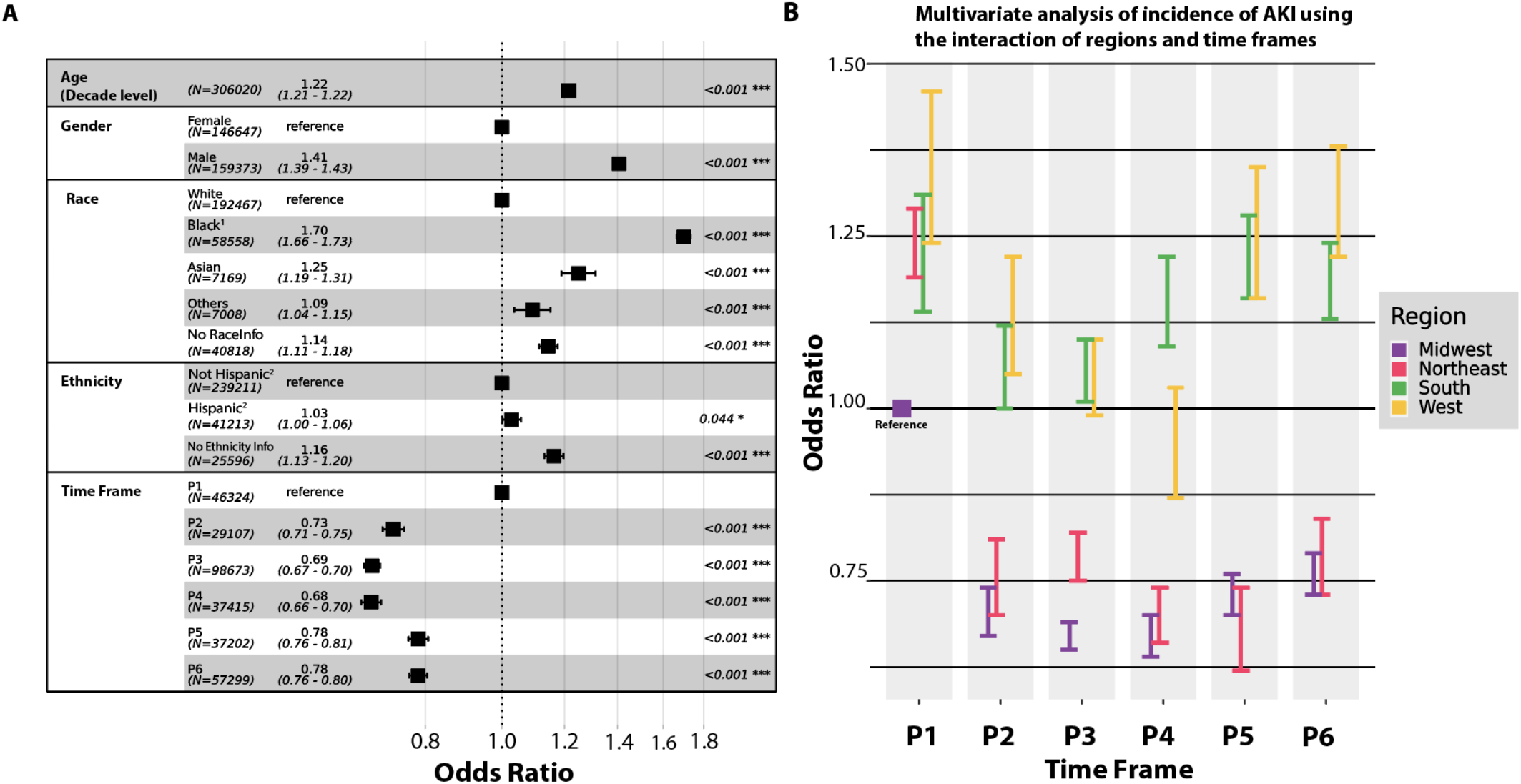
Multivariable analysis of AKI incidence for 306,020 patients using the logistic regression model. (a) Model adjusted with demographics and time frames. (b) Comparison of adjusted incidence of AKI among 24 groups based on patients in Midwest P1 (4 regions * 6 time frames)

**Figure 5.**
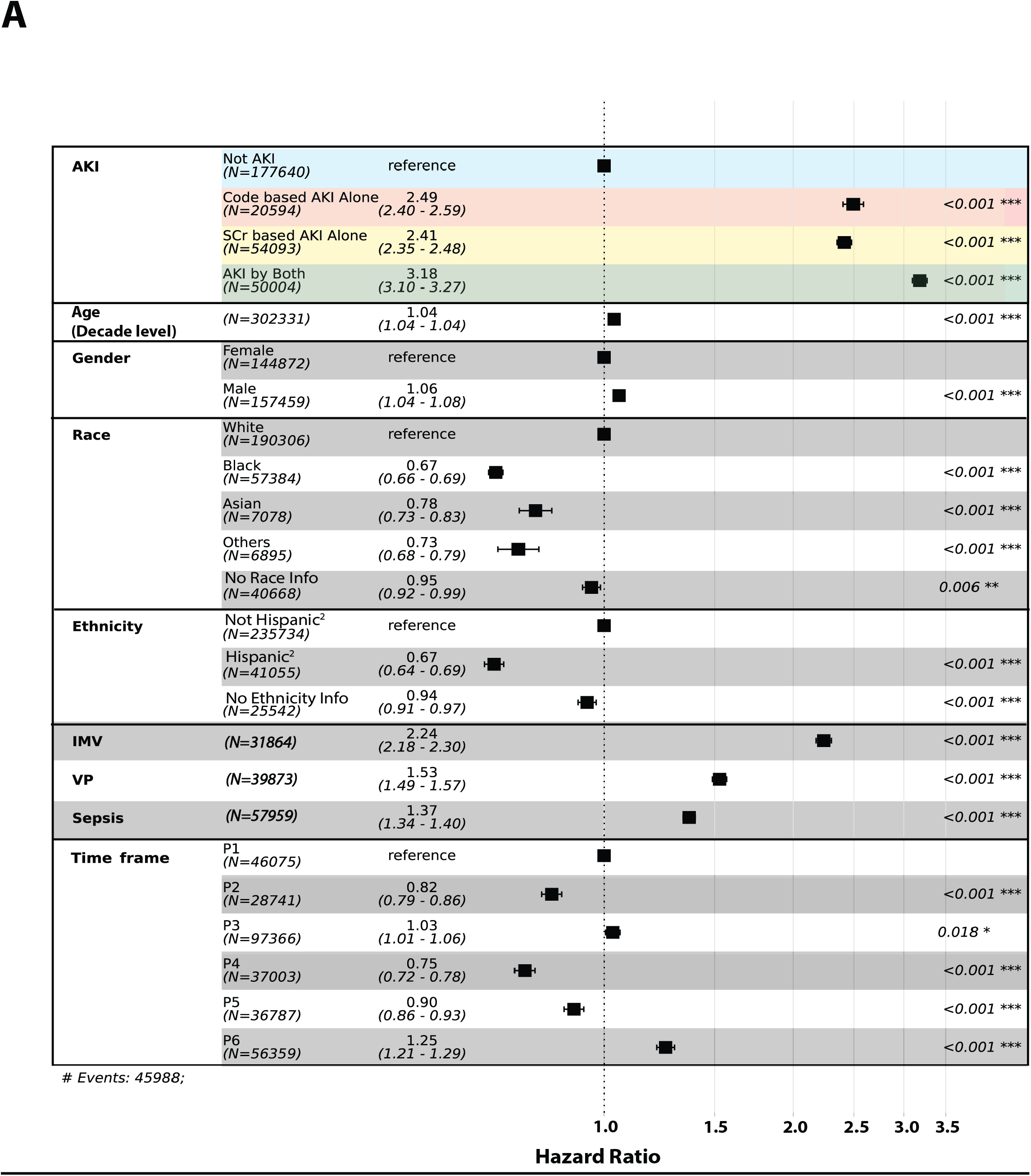
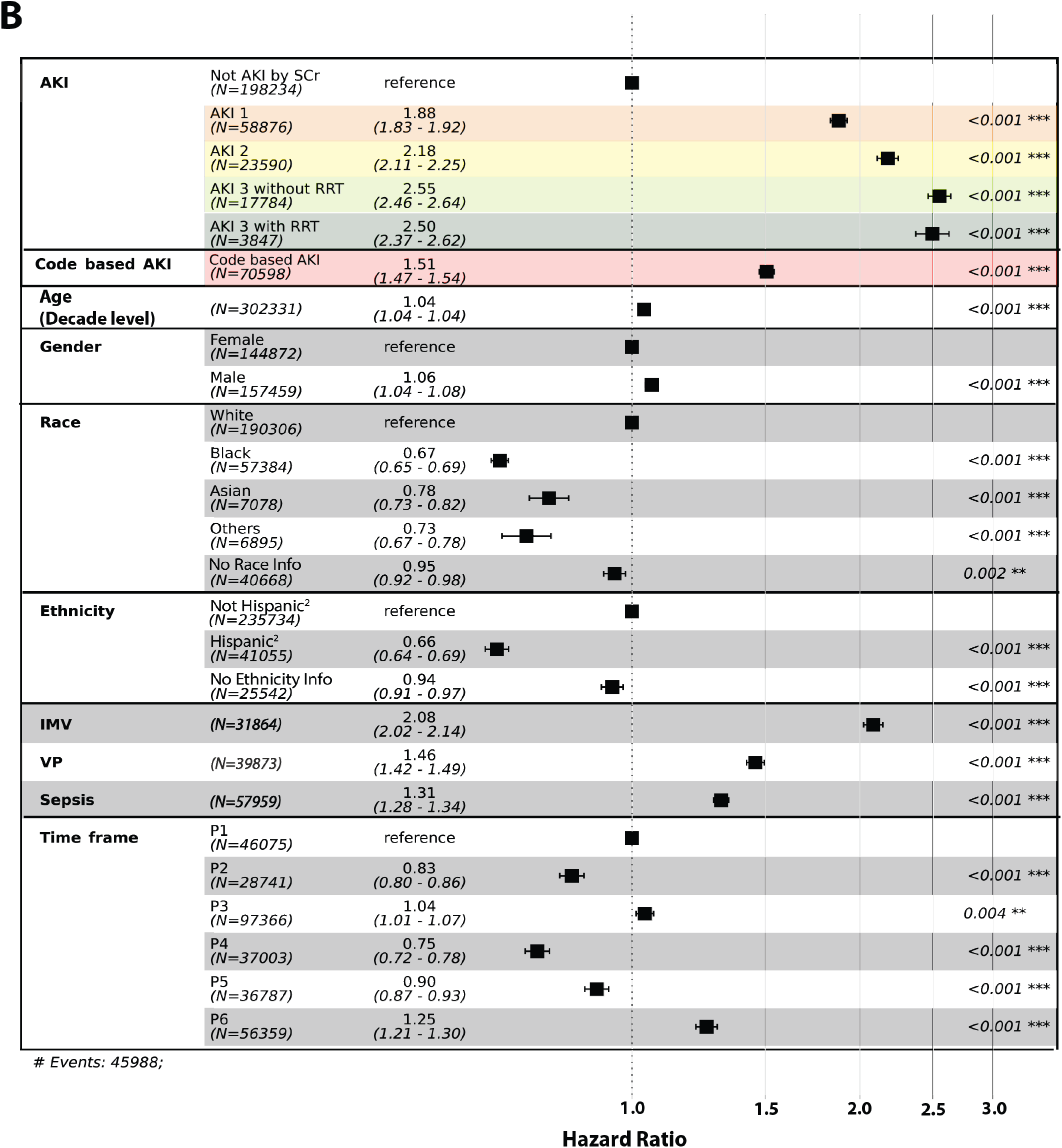
Multivariate survival analysis of 302,331 patients using the Cox proportional hazards model (same color scheme as Figures 2 and 3) A. Model including AKI by definition (Red: Code-based alone, Gold: SCr-based alone, Green: AKI by both). B. Model including Code-based alone, SCr-based AKI by Severity (Orange: AKI 1, Yellow: AKI 2, Light green: AKI 3 with RRT, Dark green: AKI 3 without RRT, Red: Code-based AKI)

## Results

### Characterization of COVID-19-associated AKI

The study cohort included 306,061 adults (age: mean 61.7 years, standard deviation [SD] 18.8; 47.9% female and 62.9% White) (**Figure 1, Table 1**). 126,478 (41.0 %) patients had AKI by either criterion (**Table 1**). 71,764 (23.4%) patients had ‘*Code-based AKI*’, and 105,603 (34.5%) had ‘*SCr-based AKI*’ and 50,889 (16.6%) met both criteria (‘AKI by both’) (**Supplementary Table 2**). **Figure 2A** shows the frequencies of overlapping criteria defining AKI, and **Supplementary Table 2** shows the characteristics of these subpopulations. Furthermore, 20,875 patients (6.8%) only met the *Code-based AKI* criteria, and 54,714 (17.9%) only met the *SCr-based AKI* criteria (**Figure 2A)**.

**Figure 2B** shows the in-hospital trajectory of SCr in study patients (except those who underwent RRT) during hospitalization. The initial median SCr levels in patients with only *code-based AKI* was significantly higher than those without AKI (**Figure 2B**). The 60-day mortality rates of patients with either *SCr-based* or *code-based AKI* was significantly higher than patients without AKI (P < 0.0001) but similar to each other (**Figure 2C**). Mortality was the highest in those who met both AKI criteria. A similar trend was noted in mortality after 60 days from hospitalization (**Supplementary Figure 2B**). The mortality rate increased with a higher AKI stage (**Supplementary Figure 2C &D**).

Most AKI cases were diagnosed within the first 5 days of hospitalization reflecting a close proximity to the COVID-19 diagnosis (**Supplementary Figure 1B**). Compared to AKI by SCr-based criteria only, the cohort with AKI by both criteria had a significantly higher proportion of more severe AKI (Stages 2 & 3) (**Supplementary Figure 1C**). 633 patients (1.2%) who received RRT were not coded for AKI (**Supplementary Figure 1C**).

In a secondary analysis of patients with a *code-based AKI* diagnosis but with <2 SCr measurements (**Figure 2A)**, the single SCr measure (when available) was higher than the admission SCr measure of those with ≥2 SCr (**Figure 2B**). Approximately 30% of these patients died within 20 days of hospitalization (**Supplementary Figure 2A**).

### Temporal and Regional Trends of COVID-19-associated AKI and mortality

The overall incidence and severity of AKI were higher in P1 and gradually decreased until July 2020 (**Figure 3**). The highest AKI rates were noted in the Northeast during P1 (first wave, spring 2020), while in other regions the highest rates were noted during the pandemic waves in the winters of 2020 and 2021 (**Figure 3A**).

The number of deaths in each time period correlated with the rates of COVID-19 hospitalization in each region of the US (**Supplementary Figure 3A**). The unadjusted mortality rates among patients with all stages of AKI saw peaks in P1, P3 and P5 periods (**Supplementary Figure 3B**).

Patients in the Northeast region were older and most likely to be non-White (**Supplementary Table 3**). The West had the highest proportion of Hispanic patients, while the South had the highest proportions of Black patients and those with comorbid conditions (cardiovascular disease [CVD], diabetes mellitus [DM], heart failure [HF] and hypertension [HTN]). The death rate was overall the lowest in the Midwest.

When comparing time periods, the highest death rate and the oldest mean age were in the P1 and P3 time periods (**Supplementary Table 4**). The highest relative numbers of males, non-Whites and Hispanics were in P1. The relative frequency of all comorbid diseases was the highest in P3.

### Risk Factors of Development of COVID-19-associated AKI

Compared to patients without AKI, those with AKI were more likely to be older, male, non-White, have greater comorbidity burden and greater severity of illness (**Table 1**). In a multivariable model based on the primary cohort of 306,020 patients (**Figure 1**), older age, male gender, non-White races and P1 time period were positive association on increased odds of AKI (**Figure 4A**). In P1, all other regions had higher adjusted AKI odds compared to P1 in the Midwest. (**Figure 4B**). After P1, the odds of AKI was higher in the South and West compared to the Northeast and Midwest with the rise in AKI noted in the recent P5 and P6 time periods.

In a multivariable analysis of AKI incidence in a sub-group of 148,703 patients with BMI and comorbidity data (**Supplementary Figure 4**), DM, HTN and HF were associated with greater risk of AKI. Morbid obesity (BMI> 40 kg/m^2^) was associated with higher, while underweight (BMI < 18.5 kg/m^2^) with lower AKI risk.

### Risk Factors of Mortality in Patients Hospitalized with COVID-19

In a multivariate analysis of mortality, AKI defined by both definitions, SCr-based AKI alone and Code-based AKI alone were all strongly associated with increased hazards compared to patients without AKI (**Figure 5A**). Furthermore, mortality risk increased with the severity of AKI (**Figure 5B**). Older age, male gender and all indicators of COVID-19 severity (sepsis diagnosis and invasive mechanical ventilation [IMV]/vasopressor use) were associated with increased hazards. Non-White races compared to Whites, and Hispanics compared to non-Hispanics, were showing a negative association with hazards. (**Figure 5**). Compared to P1, P3 & P6 time periods had positive, while P2, P4 & P5 had negative association on hazards.

In a secondary multivariable model using a sub-cohort of 148,703 patients with complete comorbidity and BMI data (**Supplementary Figure 5A**), the associations of hazards were similar, except that time period P5 had positive correlation with hazards compared to P1. While being underweight (BMI < 18.5 kg/m^2^) increases risk, all categories of high BMI are negatively associated with adjusted risk compared to normal BMI. While HF and CVD were associated with hazards, HTN was significantly associated with lower adjusted risk, and DM showed no association. Since >79% of hypertensive patients in this sub-cohort had a diagnosis of DM, CVD, or HF (**Supplementary Figure 6A)**, another model was constructed after removal of these co-morbidities (**Supplementary Figure 5B**) where HTN was no longer associated with lower mortality.

In univariate analysis (**Supplementary Table 4**), White patients were more likely to be older and have a higher proportion of CVD compared to non-Whites. No difference in follow-up time periods among the races was observed (**Supplementary Figure 6B**),

## Discussion

We report the prevalence and significance of COVID-19-associated AKI and demonstrate geographic and temporal trends over 2 years since the start of the pandemic across the entire US. N3C represents the largest and most diverse patient-level EHR data set available for public use, allowing unprecedented characterization of COVID-19-associated kidney disease and related outcomes. We used unbiased data from various EHRs that underwent a series of comprehensive and rigorous data quality controls and harmonization before analysis. Altogether, this effort enabled us to rigorously and authoritatively evaluate AKI incidence, trends, risk factors, and mortality in patients hospitalized with COVID-19.

Compared to published reports using only diagnostic codes, studies using a SCr based definition reported a higher incidence of AKI^8,25^. In our study, 17.9% of cases met the SCr-based definition, but were not coded for AKI. These patients had lower SCr on admission, and almost 70% had only stage 1 AKI, hence it is likely that mild AKI was genuinely uncoded. It is also possible, that patients were ‘clinically’ diagnosed with AKI, but these diagnoses were not accurately captured as billing codes. Alternatively, some patients may have received acute dialysis (considered ‘AKI-3’ in this study) for indications other than AKI e.g. volume overload, drug toxicity, poisoning, and did not otherwise have evidence of AKI. Uncoded AKI cases had a similar association with both short and long-term mortality as those coded for AKI, and significantly higher mortality than those without AKI, thereby confirming its clinical significance. Interestingly, patients who had AKI diagnosed by both criteria had lower survival than those diagnosed with either criterion alone, suggesting that most moderate or severe AKI cases were indeed coded. In contrast, we observed that almost 6.8% of cases were coded for AKI but not identified by the SCr criteria. We speculate that many of these patients were coded for AKI in the hospital by clinicians using information not available in our dataset, for example, urine output or baseline SCr obtained through other resources. This data emphasizes the need for greater vigilance in recognizing mild cases of AKI even if the peak SCr is within the ‘normal range’ (for example a rise in SCr from 0.6 to 1.0 mg/dl). We propose that future EHR-based studies of COVID-19-associated with AKI use both criteria of AKI diagnosis to capture the maximum number of clinically significant cases.

We observed that the incidence and severity of AKI, as well as mortality, decreased after P1 (first wave of the pandemic in the US). This might be related to the characteristics of the patients within this time period (P1) i.e., a greater relative proportion of males, non-Whites and Hispanics and greater severity of illness compared to the subsequent time periods. This observation might also reflect improved management of COVID-19, changes in virulence of SARS-CoV-2 or changing case-mix of hospitalized patients, however this needs to be determined in future studies. While the initiation of mass vaccinations since December 2020 might have played a role, the incidence and severity of AKI were already lower in the P2 period, prior to approval of COVID-19 vaccines in the US. A slight increase in adjusted AKI incidence was noted in the recent P5 and P6 time periods, where we saw the emergence of the *delta* and *omicron* variants and COVID-19 hospitalizations consisting mostly of un-vaccinated or immunocompromised patients.

The spikes in AKI cases in each geographical region of the US correlated with waves of COVID-19 in each region in the last 2 years. Compared to the Midwest, the Northeast, South and West had higher adjusted AKI incidence in P1, subsequently the South and West regions continued to have the highest relative incidence. These observations might also reflect the patients’ characteristics. South had the highest portion of Black patients and those with comorbidities. The West had the highest proportion of males and Hispanics. Both regions had patients with greater severity of illness compared to the Northeast and Midwest. It is possible that other regional differences like the type of medical centers included in the N3C database, rates of vaccinations and prevalence of specific SARS-coV-2 variants might have also played a role in the geographical variations in rates of AKI.

In multivariable models, we observed that that established risk factors like older age, male gender and Black race were associated with AKI in patients with COVID-19. In a subgroup analysis, DM, HTN, HF and morbid obesity were also associated with AKI. AKI was highly associated with mortality and higher stage of AKI was associated with a greater risk of death. Consistent with previous studies^26^, older age, male gender and severity of illness were associated with higher mortality. An unexpected finding in our multivariable models for morality was that non-White race and Hispanic ethnicity was associated with a lower adjusted mortality risk similar to findings from a recent study^27^. However, these observations should be interpreted in light of the limitations of this study. Importantly, a significant number of patients in our cohort had no information on race (13.34%) or ethnicity (8.37%). Also, our cohort consisted of only hospitalized patients, and we do not have data on AKI incidence and mortality prior to index hospitalization or in those who were never hospitalized. It is possible that Black and Hispanic patients with less access to care had a higher out-of-hospital mortality or that many were hospitalized in centers not part of the N3C, and therefore not represented in our data.

In multivariate models for a subgroup of patients with complete medical history and BMI, unexpectedly, those with HTN appeared to have a lower mortality risk, unlike other studies of patients with COVID-19 where HTN was associated with death^28,29^. However in multivariate models excluding more severe comorbidities (DM, HF and CVD), HTN was no longer associated with mortality. While we also observed the previously reported J-shape mortality curve with BMI categories^30^, in our study, obesity was associated with lower adjusted mortality compared to those with normal BMI. The association of obesity with mortality is variable in observational studies^31-34^. Again, these HTN and BMI associations should be interpreted with caution since almost half of our cohort was excluded in these sub-group analyses.

Our study has several limitations in addition to those mentioned above. This is an observational study; hence causality cannot be determined. While we controlled for the major risk factors for AKI and mortality in our multivariable models, residual confounding due to other factors like laboratory biomarkers and medications is possible. We show that using both SCr-based and code-based criteria for AKI diagnosis reduces false-negative cases of AKI, however, we might have missed a significant number of AKI cases due to the lack of data on urine output. While data from all geographical regions of the US is represented, there is a relative lack of data from non-academic medical centers. N3C data is a diverse but not necessarily representative mixture of individual patient experiences, risks, providers, and system practice patterns, as viewed through the lens of EHR-based data ingestion and harmonization processes to the N3C-employed OMOP common data model. An ontology-based data annotation and standardization approach can enhance the data integration and analysis^35^. Additional survival analyses that modeled inter-site heterogeneity did not alter findings. Our study also did not include data from other countries that may show different patients and viral characteristics^36^, hence our findings might not be reflective of COVID-19-associated AKI patterns in other world regions. We are also likely missing many deaths occurring after hospital discharge. At the time of this study, we did not have reliably complete data on vaccinations.

In conclusion, we report that the incidence of COVID-19-asssociated AKI has reduced after the first wave of the pandemic in USA and regional differences in AKI and mortality rates were observed. Importantly, COVID-19-associated AKI measured by rise in SCr, even if not coded for by clinicians during hospitalization, was significantly associated with mortality. This observation suggests that greater attention to the presence of AKI is warranted in the setting of hospitalization for COVID-19, to potentially help with improvements in management and outcomes. We recommend future studies use more than one criteria for AKI identification.

## Supporting information

STROBE checklist

## Data Availability

All data produced in the present study are available in the N3C data enclave with approval from NCATS.

## Acknowledgements

The primary study sponsors are multiple institutes of the National Institutes of Health. The NCATS is the primary steward of the N3C data, created the underlying architecture of the N3C Data Enclave (covid.cd2h.org/enclave), manages the Data Transfer Agreements and Data Use Agreements, houses the Data Access Committee, and supports contracts to vendors to help build various aspects of the N3C Data Enclave. This research was possible because of the patients whose information is included within the data from participating organizations (covid.cd2h.org/dtas) and the organizations and scientists (covid.cd2h.org/duas) who have contributed to the on-going development of this community resource^15^. The content is solely the responsibility of the authors and does not necessarily represent the official views of the NIH. The analyses described in this publication were conducted with data or tools accessed through the NCATS N3C Data Enclave and supported by NCATS U24 TR002306. Authorship was determined using ICMJE recommendations.

## Data Partners with Released Data

Stony Brook University — U24TR002306 • University of Oklahoma Health Sciences Center — U54GM104938: Oklahoma Clinical and Translational Science Institute (OCTSI) • West Virginia University — U54GM104942: West Virginia Clinical and Translational Science Institute (WVCTSI) • University of Mississippi Medical Center U54GM115428: Mississippi Center for Clinical and Translational Research (CCTR) • University of Nebraska Medical Center — U54GM115458: Great Plains IDeA-Clinical & Translational Research • Maine Medical Center U54GM115516: Northern New England Clinical & Translational Research (NNE-CTR) Network • Wake Forest University Health Sciences — UL1TR001420: Wake Forest Clinical and Translational Science Institute • Northwestern University at Chicago — UL1TR001422: Northwestern University Clinical and Translational Science Institute (NUCATS) • University of Cincinnati — UL1TR001425: Center for Clinical and Translational Science and Training • The University of Texas Medical Branch at Galveston — UL1TR001439: The Institute for Translational Sciences • Medical University of South Carolina — UL1TR001450: South Carolina Clinical & Translational Research Institute (SCTR) • University of Massachusetts Medical School Worcester — UL1TR001453: The UMass Center for Clinical and Translational Science (UMCCTS) • University of Southern California — UL1TR001855: The Southern California Clinical and Translational Science Institute (SC CTSI) • Columbia University Irving Medical Center — UL1TR001873: Irving Institute for Clinical and Translational Research • George Washington Children’s Research Institute — UL1TR001876: Clinical and Translational Science Institute at Children’s National (CTSA-CN) • University of Kentucky — UL1TR001998: UK Center for Clinical and Translational Science • University of Rochester — UL1TR002001: UR Clinical & Translational Science Institute • University of Illinois at Chicago — UL1TR002003: UIC Center for Clinical and Translational Science • Penn State Health Milton S. Hershey Medical Center — UL1TR002014: Penn State Clinical and Translational Science Institute • The University of Michigan at Ann Arbor — UL1TR002240: Michigan Institute for Clinical and Health Research • Vanderbilt University Medical Center — UL1TR002243: Vanderbilt Institute for Clinical and Translational Research • University of Washington — UL1TR002319: Institute of Translational Health Sciences • Washington University in St. Louis — UL1TR002345: Institute of Clinical and Translational Sciences • Oregon Health & Science University — UL1TR002369: Oregon Clinical and Translational Research Institute • University of Wisconsin-Madison — UL1TR002373: UW Institute for Clinical and Translational Research • Rush University Medical Center — UL1TR002389: The Institute for Translational Medicine (ITM) • The University of Chicago — UL1TR002389: The Institute for Translational Medicine (ITM) • University of North Carolina at Chapel Hill — UL1TR002489: North Carolina Translational and Clinical Science Institute • University of Minnesota — UL1TR002494: Clinical and Translational Science Institute • Children’s Hospital Colorado — UL1TR002535: Colorado Clinical and Translational Sciences Institute • The University of Iowa — UL1TR002537: Institute for Clinical and Translational Science • The University of Utah — UL1TR002538: Uhealth Center for Clinical and Translational Science • Tufts Medical Center — UL1TR002544: Tufts Clinical and Translational Science Institute • Duke University — UL1TR002553: Duke Clinical and Translational Science Institute • Virginia Commonwealth University — UL1TR002649: C. Kenneth and Dianne Wright Center for Clinical and Translational Research • The Ohio State University — UL1TR002733: Center for Clinical and Translational Science • The University of Miami Leonard M. Miller School of Medicine — UL1TR002736: University of Miami Clinical and Translational Science Institute • University of Virginia — UL1TR003015: iTHRIV Integrated Translational health Research Institute of Virginia • Carilion Clinic — UL1TR003015: iTHRIV Integrated Translational health Research Institute of Virginia • University of Alabama at Birmingham — UL1TR003096: Center for Clinical and Translational Science • Johns Hopkins University — UL1TR003098: Johns Hopkins Institute for Clinical and Translational Research • University of Arkansas for Medical Sciences — UL1TR003107: UAMS Translational Research Institute • Nemours — U54GM104941: Delaware CTR ACCEL Program • University Medical Center New Orleans U54GM104940: Louisiana Clinical and Translational Science (LA CaTS) Center • University of Colorado Denver, Anschutz Medical Campus — UL1TR002535: Colorado Clinical and Translational Sciences Institute • Mayo Clinic Rochester — UL1TR002377: Mayo Clinic Center for Clinical and Translational Science (CCaTS) • Tulane University — UL1TR003096: Center for Clinical and Translational Science • Loyola University Medical Center — UL1TR002389: The Institute for Translational Medicine (ITM) • Advocate Health Care Network — UL1TR002389: The Institute for Translational Medicine (ITM) • OCHIN — INV-018455: Bill and Melinda Gates Foundation grant to Sage Bionetworks

## Article Information

### Corresponding Authors

Richard A. Moffitt, PhD, Farrukh M. Koraishy, MD, PhD.

### Author Contributions

Dr.Moffitt and Dr.Koraishy have full access to all data in the study and are responsible for the integrity of the data and the accuracy of data analysis.

*Concept and design*: Yun Jae Yoo, Kenneth J. Wilkins, Spencer Krichevsky, Stephanie S. Hong, Feifan Liu, Chetan K. Potu, Richard L. Zhu, Luke A. Torre-Healy, Rajiv Saran, Yongqun He, Fadhl Alakwaa, Ankit Sakhuja, Joel H. Saltz, Soko Setoguchi, Sandra L. Kane-Gill, Sandeep K. Mallipattu, David H. Ellison, James Brian Byrd, Chirag R. Parikh, Richard A. Moffitt and Farrukh M. Koraishy

*Acquisition, analysis, or interpretation of data*: All authors.

*Drafting of the manuscript*: Yoo, Krichevsky, Moffitt, Koraishy.

*Critical revision of the manuscript for important intellectual content*: All authors.

*Statistical analysis*: Yoo, Moffitt, Wilkins, Setoguchi.

*Administrative, technical, or material support*: Consortial authors

*Supervision*: Moffitt, Koraishy.

## Additional Contributions

Tellen Bennett, Katie Bradwell, Christopher Chute, Peter DeWitt, Andrew Girvin, Davera Gabriel, Janos Hajagos, Melissa Haendel, Harold Lehmann, Emily Pfaff, Siao Sun, Jacob Wooldridge.

## Figure legends

**Figure 1. Flow diagram showing the total number of patients used in the study and the patients used in each table and figure**

The study included all adult patients in the N3C data set with COVID-19 positive within 7 days of an inpatient hospitalization. Other patients were excluded for study due to site-specific data quality issues, pre-existing ESRD, or lack of SCr data.

**Figure 2. Development of the cohort, definitions of COVID-19-associated AKI and comparative mortality rates.**

(a) Overall prevalence of AKI by different AKI definitions in a total of 306,061 adult cohorts. (b) SCr trajectories by day of admission between patient groups classified in different AKI definitions. (c) 60-day survival after diagnosis of COVID-19 in the patient shown in a.

**Figure 3. Temporal and geographical distribution of COVID-19-associated AKI**

(a) Timeline of patients with and without AKI across four regions of the United States of our cohort of 306,061. (b) Choropleths show the overall number of the patients and sites over region. (c) Timeline of definitions of AKI among those with AKI over time shows a higher level of more AKI by both at earlier times. (d) Timeline of severity of AKI among those with AKI over time shows a higher level of more severe disease at earlier times.

**Figure 4. Multivariable analysis of AKI incidence for 306,020 patients using the logistic regression model**

(a) Model adjusted with demographics and time frames.

(b) Comparison of adjusted incidence of AKI among 24 groups based on patients in Midwest P1 (4 regions * 6 time frames)

**Figure 5. Multivariate survival analysis of 302,331 patients using the Cox proportional hazards model (same color scheme as Figures 2 and 3)**

(a) Model including AKI by definition (Red: Code-based alone, Gold: SCr-based alone, Green: AKI by both).

(b) Model including Code-based alone, SCr-based AKI by Severity (Orange: AKI 1, Yellow: AKI 2, Light green: AKI 3 with RRT, Dark green: AKI 3 without RRT, Red: Code-based AKI)

