## Supplemental methods, tables, and figures for "COVID-19-associated AKI in hospitalized US patients: incidence, temporal trends, geographical distribution, risk factors and mortality"

**Supplementary Appendix**.

***Supplemental Online Contents***

#### Supplementary Methods

#### Supplementary Tables

**Supplementary Table 1:** The defined list of concepts including visit type, race, ethnicity, comorbidities (AKI, DM, HTN, CKD, HF, ESRD, Sepsis), procedures (ECMO, IMV) and drugs (ACEIs, ARBs, VP).
**Supplementary Table 2:** Descriptive characteristics of hospitalized COVID positive patients with and without AKI by two different AKI definitions.
**Supplementary Table 3:** Descriptive characteristics of each region
**Supplementary Table 4:** Descriptive characteristics of each time period
**Supplementary Table 5:** Descriptive characteristics of each racial group.

#### Supplementary Figures:

**Supplementary Figure 1:** (a) Venn diagrams showing patients meeting different SCr-based AKI definitions **(b)** Comparison between the time of onset of AKI from the date of hospitalization and the length of hospitalization of patients (code-based AKI for all patients with AKI onset longer than the length of hospitalization). Color means the number of patients, darker the more patients. (**c)** Comparison of severity between patients meeting both AKI criteria *versus* those meeting only SCr criteria.
**Supplementary Figure 2: (a)** First 60-day survival for patients diagnosed with AKI, but with insufficient SCr data to calculate a change. **(b)** Post-60-day survival for different AKI definitions. **(c)** First 60-day survival by KDIGO-based AKI stages and Code-based AKI. **(d)** Post-60-day survival by KDIGO-based AKI stages and Code-based AKI.
**Supplementary Figure 3: (a)** Comparison of univariate mortality rates (and 95% confidence intervals), between regional ad time frames. **(b)** Observed mortality rates (and 95% confidence intervals), within different severities of AKI, for six time periods.
**Supplementary Figure 4:** Multivariate analysis of COVID-19-related AKI risk in patients with BMI
**Supplementary Figure 5: (a)** Multivariable survival analysis of 148,703 patients using the Cox Proportional Hazards (CoxPH) model BMI and comorbidity. **(b)** CoxPH multivariate analysis including only HTN with BMI.
**Supplementary Figure 6: (a)** Follow-up comparison between Race groups. (b) Venn-diagram of Comorbidities (HTN, DM, HF, CVD).

**Supplemental Methods**

#### Variable Definitions

Visit types

Relevant inpatient visits in the OMOP format are described by five visit-type concepts. To unite these terms semantically, we created concept sets defining not only inpatient visits, but also emergency department (ED) visits and outpatient visits (**Supplementary Table 1**). Furthermore, visits for a single patient of any type overlapping in time with inpatient visits were merged using the 'macrovisit' logic described previously[1].

Dates, survival, and length of stay

To construct length of follow-up, i.e., censoring for survival analysis, we define the date of last follow-up as each patient's most recent measurement date. The date of death was not available for 411 of 46,122 (<1%) deceased patients. When missing, the date of death was assumed to be the date of the last measurement data available. When the date of death or last follow-up was known to be after the study end date, patients were treated as being alive at the study end date and censored accordingly (n=3,166; administrative censoring).

Race and Ethnicity

Data on race and ethnicity were obtained from the EHR, which may represent a mixture of self-reported and observed information. Heterogeneous OMOP concepts for race were combined into five racial groups as described in **Supplementary Table 1**.

*Covariates*

Conditions, medications, and laboratory values were defined using concept sets, i.e., sets of semantically similar OMOP concepts, that are freely available for re-use within the N3C enclave. Many concept sets have been previously defined and validated[1], but additional concept sets were built for this study (**Supplementary Table 1**). For laboratory values, all data were converted to a single unit of measurement prior to computation.

Inclusion and Exclusion Criteria

The following site-level data quality criteria was also applied to the cohort (**Figure 1**).

1) *Long-term facilities*. We calculated the date difference between the date of the confirmed case of COVID-19 and the start date of the visit to determine the proportion of inpatients at each hospital with a date difference of more than 200 days (indicating a hospital-acquired infection). If the number of these long-term inpatients exceeded 5% of all inpatients, we classified the facility as 'long-term' and excluded such sites and their patients (n=1 site and n=6,417 patients) from the study **(Figure 1)**.
2) *Availability of serum creatinine (SCr) data*. Preliminary analysis of the N3C data suggested systematic missingness of SCr data by site, and that while most sites had excellent (>90%) coverage of SCr amongst inpatients, 3 sites were removed because no SCr data were available, and 15 additional sites were removed for providing SCr from fewer than <75% of inpatients **(Figure 1)**.
3) *Availability of mortality data*. While most sites reported COVID-19-positive inpatient mortality above 8% (range of 4-23%), we dropped 1 site from survival endpoint studies that reported mortality below 3% **(Figure 1)**.

**Renal Measures:**

*Definition of baseline serum creatinine (SCr)*

A patient's 'baseline' SCr was established using the following procedure, with the most recent pre-hospitalization SCr taking precedence if available in the last 180 days prior to the COVID visit: (1) If a patient had an outpatient SCr before their COVID-19 inpatient visit (index visit), we selected the most recent outpatient SCr value as a baseline for that patient. (2) If the patient did not have an outpatient SCr prior to the index visit but had an emergency department (ED) visit SCr prior to the COVID-19 visit, we selected the SCr at discharge from the ED visit as the patient's baseline. (3) If a patient had neither an outpatient nor an ED visit SCr, but they had an inpatient SCr before their index visit, we selected the SCr at discharge for that inpatient visit. (4) If a patient did not have any SCr before the index visit, we selected the minimum SCr of all the SCr obtained during the index hospitalization as reported previously[2,3]. SCr values provided by different institutions or labs were harmonized to the same unit of measure (mg/dL)[4].

*Definition of AKI severity*

The severity of AKI was defined as follows. First, a common feature is the use of a set of concepts for renal replacement therapy (RRT), which defined all patients who received RRT or acute dialysis during the COVID-19 visit as “AKI stage 3 with RRT”. Among patients with "SCr-based AKI”, Stage 1 AKI was defined as a SCr fold-change of at least 1.5 but less than 2 from baseline, Stage 2 was defined as a change of at least 2-fold, but less than 3-fold. Stage 3 AKI without RRT was defined as SCr fold change of at least 3 times the baseline but not requiring acute dialysis. In the case of an increase in SCr of 0.3 or more within 48 hours, all differences greater than 0.3 were defined as AKI stage 1. When the same patient showed different AKI stages in different SCr-based AKI definitions, the severity of the patients was defined by selecting the highest stage.

#### BMI Interpolation

BMI, height, and weight values were not consistently reported on the patient- nor site-level. For example, 8 of 53 (15%) sites were unable to transmit BMI data and 148,722 of 306,061 (49%) patients had ≥1 reported BMI value. To enrich the dataset and facilitate upstream analyses, a workflow was implemented to mitigate the degree at which BMI measurements were seemingly missing throughout the study cohort. Missing weight and height data were interpolated by inserting the most recent validated measurement occurring within seven days or one year, respectively. These criteria were pre-defined to ensure that interpolated values exhibited stability and to minimize the potential for data corruption as a source of bias. The aforementioned process by which SCr units and other metrics were harmonized was also applied to these data. Missing BMI measurements were then calculated according to the following formula: $BMI = weight\ [kg] / (height\ [m])^2$. This workflow decreased the number of sites without any BMI data to 1/53 (2%), representing an 88% reduction in the number of sites missing this established COVID-19 risk factor. Additionally, the number of sites with >95% reported BMI data increased from

1/53 (2%) to 43/53 (81%) and the number of patients with ≥1 reported BMI value increased to 202972/306061 (66%).

Statistical Analysis

The AKI incidence probability is subdivided into regions and times, and the adjusted odds ratio (OR) values are shown based on basic demographic information (**Figure 4B**). Multivariable Cox proportional hazards (CoxPH) regression was used to estimate hazard ratios (HRs) for survival endpoints (**Figure 5**). Primary multivariable analyses were performed based on 306,020 patients, excluding 41 patients (0.01%) without gender information (**Figure 1**). There were no missing values in any of the primary multi-variable models that were designed using variables with complete data on the entire cohort: age [by decade], gender, race, ethnicity and observation time periods (P1, P2, P3, P4, P5, P6) (**Figures 4 and 5**) and for mortality prediction, the variables associated with severity of illness during hospitalization (the need for Invasive Mechanical Ventilation [IMV] or vasopressors [VP] use or diagnosis of sepsis) were also included (**Figure 5**).

Secondary multivariable analysis was done on a sub-group of patients with data on comorbidities and body mass index (BMI) (**Figure 1**). Comorbidities included hypertension (HTN), diabetes (DM), heart failure (HF), and cardiovascular disease (CVD). Secondary Multivariate analyses are shown in **Supplementary figures 4 and 5 and** included 148,703 patients, after excluding 157,358 patients without BMI records.

Follow-up visit analyses in **Supplementary Figure 6** were based on Kaplan-Meier curves from the *survival* package in R[5]. The start date was the date of confirmed COVID-19, and the end date was the most recent date with measurement data. The design of N3C limits the potential to adjust for covariates that are not directly measurable. However, allowing for data-partner heterogeneity within survival analyses serves as a tractable yet informal assessment of how conclusions may be impacted by such latent factors in aggregate. Survival analyses were repeated using shared frailty models (as done for time-to-event analyses in other multi-site studies of COVID-19)[6,7] for assessing robustness of the primary Cox regression model and data-partner-specific Kaplan-Meier survival curves (not shown) to affirm that primary survival analyses' conclusions were similar. The patients included in the analysis were counted only once based on the start date of their index COVID-19 hospitalization.

***Supplementary Tables***

**Supplementary Table 1: The defined list of concepts includes visit type, race, ethnicity, comorbidities (AKI, DM, HTN, CKD, HF, ESRD, sepsis), procedures (RRT, IMV) and drugs (VP).**

| | |
|---|---|
| **Visit Type** | **Concept name** |
| **Inpatient** | |
| | inpatient visit |
| | inpatient hospital |
| | emergency room and inpatient visit |
| | inpatient intensive care unit |
| | inpatient psychiatric facility |
| **Visit Type** | **Concept name** |
| **Outpatient** | |
| | Outpatient Visit |

| | |
|---|---|
| | Office Visit |
| | Laboratory Visit |
| | Observation Room |
| | Non-hospital institution Visit |
| | Ambulatory Surgical Center |
| | Ambulatory Clinic / Center, Ambulatory Infusion Therapy Clinic / Center |
| | Interactive Telemedicine Service |
| | Telehealth |
| | Ambulatory Oncology Clinic / Center, Ambulatory Radiology Clinic / Center |
| | Telephone call to a patient |
| | Ambulatory Endoscopy Clinic / Center |
| | Ambulatory Oncological Radiation Clinic / Center |
| | Ambulatory Dental Clinic / Center |
| | Ambulatory Magnetic Resonance Imaging (MRI) Clinic / Center |
| | Skilled Nursing Facility |
| **Race** | **Concept name** |
| **White** | |
| | White |
| **Black** | |
| | Black |
| | Black or African American |
| **Asian** | |
| | Asian |
| | Asian Indian |
| | Filipino |
| | Korean |
| | Chinese |
| | Vietnamese |
| | Japanese |
| **Others** | |
| | other |
| | different races |
| | multiple races |
| | two or more races |
| | Native Hawaiian or other Pacific Islander |
| | Other Pacific Islander |
| | Polynesian |
| | Hispanic |

| | |
|---|---|
| **Ethnicity** | |
| **Not Hispanic or Latino Ethnicity** | |
| | Not Hispanic or Latino Ethnicity |
| **Hispanic or Latino** | |
| | Hispanic or Latino |
| | **Diagnostic concept name** |
| Acute Kidney Injury (AKI) | |
| | Acute injury of kidney |
| | Acute tubular necrosis |
| | Hepatorenal syndrome |
| | Acute renal failure due to acute cortical necrosis |
| | Hemolytic uremic syndrome |
| | Acute-on-chronic renal failure |
| | Acute renal insufficiency |
| | Postpartum acute renal failure |
| | Acute nephritis |
| | Rapidly progressive nephritic syndrome |
| | Acute renal impairment |
| | Acute kidney injury due to sepsis |
| | Acute tubulointerstitial nephritis |
| | Rapidly progressive nephritic syndrome |
| | diffuse crescentic glomerulonephritis |
| | Acute nontraumatic kidney injury, |
| | Acute renal failure on dialysis |
| | Acute kidney failure stage 1-3 |
| | Rapidly progressive glomerulonephritis |
| | Hemorrhagic fever with renal syndrome |
| | Acute renal cortical necrosis |
| | Rapidly progressive nephritic syndrome |
| | diffuse membranous glomerulonephritis |
| | Acute renal failure due to tubular necrosis |
| | Acute kidney injury due to hypovolemia. |
| | **Diagnostic concept name** |

| **Diabetes (DM)** | |
|---|---|
| | Type 2 diabetes mellitus |
| | Type 2 diabetes mellitus without complication |
| | Hyperglycemia due to type 2 diabetes mellitus |
| | Chronic kidney disease due to type 2 diabetes mellitus |
| | Complication due to diabetes mellitus |
| | Polyneuropathy due to type 2 diabetes mellitus |
| | Disorder of nervous system due to type 2 diabetes mellitus |
| | Peripheral circulatory disorder due to type 2 diabetes mellitus |
| | Foot ulcer due to type 2 diabetes mellitus |
| | Renal disorder due to type 2 diabetes mellitus |
| | Type 1 diabetes mellitus |
| | Secondary diabetes mellitus |
| | Disorder due to type 2 diabetes mellitus |
| | Diabetic ketoacidosis without coma |
| | Proliferative retinopathy due to type 2 diabetes mellitus |
| | Diabetes mellitus without complication |
| | Hyperglycemia due to type 1 diabetes mellitus |
| | Disorder of eye due to type 2 diabetes mellitus |
| | Hypoglycemia due to type 2 diabetes mellitus |
| | Type 2 diabetes mellitus with ulcer |
| | Diabetes mellitus |
| | Mild nonproliferative retinopathy due to type 2 diabetes mellitus |
| | Type 1 diabetes mellitus without complication |
| | Macular edema due to diabetes mellitus |
| | Autonomic neuropathy due to type 2 diabetes mellitus |
| | Gestational diabetes mellitus |
| | Macular edema and retinopathy due to type 2 diabetes mellitus |
| | Disorder of nervous system due to diabetes mellitus |
| | Insulin treated type 2 diabetes mellitus |
| | Polyneuropathy due to diabetes mellitus |
| | Disorder of kidney due to diabetes mellitus |
| | Dermopathy due to type 2 diabetes mellitus |
| | Moderate nonproliferative retinopathy due to type 2 diabetes mellitus |
| | Drug-induced diabetes mellitus |
| | Type II diabetes mellitus uncontrolled |
| | Renal disorder due to type 1 diabetes mellitus |
| | Gangrene due to type 2 diabetes mellitus |

| | |
|---|---|
| | Peripheral angiopathy due to diabetes mellitus |
| | Pre-existing type 2 diabetes mellitus in pregnancy |
| | Cataract due to diabetes mellitus type 2 |
| | Diabetic foot ulcer |
| | Disorder of eye due to diabetes mellitus |
| | Neuropathic arthropathy due to type 2 diabetes mellitus |
| | Diabetes insipidus |
| | Hypoglycemia due to type 1 diabetes mellitus |
| | Disorder of nervous system due to type 1 diabetes mellitus |
| | Pre-existing type 1 diabetes mellitus in pregnancy |
| | Autonomic neuropathy due to type 1 diabetes mellitus |
| | Polyneuropathy due to type 1 diabetes mellitus |
| | Type 2 diabetes mellitus well controlled |
| | Nonproliferative retinopathy due to type 2 diabetes mellitus |
| | Proliferative retinopathy due to diabetes mellitus |
| | Retinopathy due to type 2 diabetes mellitus |
| | Neuropathy due to diabetes mellitus |
| | Ulcer of lower limb due to type 1 diabetes mellitus |
| | Mononeuropathy due to type 2 diabetes mellitus |
| | Neuropathy due to type 2 diabetes mellitus |
| | Severe hyperglycemia due to diabetes mellitus |
| | Type 2 diabetes mellitus with peripheral angiopathy |
| | Peripheral vascular disorder due to diabetes mellitus |
| | Severe nonproliferative retinopathy with clinically significant macular edema due to diabetes mellitus |
| | Retinopathy due to type 1 diabetes mellitus |
| | Proliferative retinopathy due to type 1 diabetes mellitus |
| | Mild nonproliferative retinopathy due to diabetes mellitus |
| | Pre-existing diabetes mellitus in pregnancy |
| | Moderate nonproliferative retinopathy due to diabetes mellitus |
| | Diabetes mellitus during pregnancy, childbirth and the puerperium |
| | Type 1 diabetes mellitus uncontrolled |
| | Ketoacidosis due to type 2 diabetes mellitus |
| | Severe nonproliferative retinopathy without macular edema due to diabetes mellitus |
| | Autonomic neuropathy due to diabetes mellitus |
| | Retinopathy due to diabetes mellitus |
| | Nonproliferative retinopathy due to diabetes mellitus |
| | Ketoacidosis due to type 1 diabetes mellitus |
| | Peripheral circulatory disorder due to type 1 diabetes mellitus |

| | |
|---|---|
| | Hyperosmolar coma due to diabetes mellitus |
| | Hyperosmolar coma due to type 2 diabetes mellitus |
| | Diabetic ketoacidosis |
| | Latent autoimmune diabetes mellitus in adult |
| | Disorder due to type 1 diabetes mellitus |
| | Severe nonproliferative retinopathy due to diabetes mellitus |
| | Lumbosacral radiculoplexus neuropathy due to type 2 diabetes mellitus |
| | Arthropathy due to type 2 diabetes mellitus |
| | Diabetes mellitus in mother complicating pregnancy, childbirth AND/OR puerperium |
| | Disorder of eye due to type 1 diabetes mellitus |
| | Mild nonproliferative retinopathy due to type 1 diabetes mellitus |
| | Type 2 diabetes mellitus in obese |
| | Gestational diabetes mellitus in childbirth |
| | O/E - right eye proliferative diabetic retinopathy |
| | Infection of foot due to diabetes mellitus |
| | Proliferative retinopathy of right eye with diabetes mellitus |
| | Type 1 diabetes mellitus with ulcer |
| | Gastroparesis due to diabetes mellitus |
| | O/E - left eye proliferative diabetic retinopathy |
| | Cataract due to diabetes mellitus type 1 |
| | Mild nonproliferative retinopathy of right eye due to diabetes mellitus |
| | Mild nonproliferative retinopathy of left eye due to diabetes mellitus |
| | Moderate nonproliferative retinopathy due to type 1 diabetes mellitus |
| | Traction detachment of retina due to type 2 diabetes mellitus |
| | Diabetic mononeuropathy |
| | Hypoglycemia due to diabetes mellitus |
| | Skin ulcer due to diabetes mellitus |
| | Nonproliferative diabetic retinopathy due to type 1 diabetes mellitus |
| | Pre-existing diabetes mellitus in mother complicating childbirth |
| | Nephrogenic diabetes insipidus |
| | Ketoacidotic coma due to type 1 diabetes mellitus |
| | Diabetic neuropathy with neurologic complication |
| | Diabetes mellitus type 2 without retinopathy |
| | Type 2 diabetes mellitus in nonobese |
| | Dermopathy due to type 1 diabetes mellitus |
| | Macular edema due to type 2 diabetes mellitus |
| | Cataract due to diabetes mellitus |
| | Ulcer of midfoot due to diabetes mellitus |

| | |
|---|---|
| | Hyperosmolar coma due to secondary diabetes mellitus |
| | Mononeuropathy due to type 1 diabetes mellitus |
| | Ulcer of toe due to type 2 diabetes mellitus |
| | Chronic kidney disease due to type 1 diabetes mellitus |
| | Moderate nonproliferative diabetic retinopathy of right eye |
| | Diabetes mellitus during pregnancy - baby not yet delivered |
| | Hypoglycemic coma due to type 2 diabetes mellitus |
| | Ketoacidotic coma due to type 2 diabetes mellitus |
| | Steroid-induced diabetes |
| | Neuropathic arthropathy due to type 1 diabetes mellitus |
| | Gestational diabetes mellitus, class A>2< |
| | Moderate nonproliferative diabetic retinopathy of left eye |
| | Dyslipidemia due to type 2 diabetes mellitus |
| | Ketoacidotic coma due to diabetes mellitus |
| | Coma due to diabetes mellitus |
| | Type 2 diabetes mellitus controlled by diet |
| | Chronic kidney disease stage 3 due to type 2 diabetes mellitus |
| | Chronic kidney disease stage 3 due to type 1 diabetes mellitus |
| | Mixed hyperlipidemia due to type 2 diabetes mellitus |
| | Gangrene due to type 1 diabetes mellitus |
| | Gestational diabetes mellitus complicating pregnancy |
| | Pregnancy and type 2 diabetes mellitus |
| | Posttransplant diabetes mellitus |
| | Pregnancy and type 1 diabetes mellitus |
| | Diabetic peripheral neuropathy |
| | Proliferative retinopathy of left eye due to diabetes mellitus |
| | Hypoglycemic coma due to type 1 diabetes mellitus |
| | Diabetic dermopathy |
| | Ulcer of heel due to diabetes mellitus |
| | Disorder due to well controlled type 2 diabetes mellitus |
| | Gestational diabetes mellitus, class A>1< |
| | Macular edema of right eye due to diabetes mellitus |
| | Diabetic foot |
| | Lumbosacral radiculoplexus neuropathy due to diabetes mellitus |
| | Proteinuric nephropathy due to diabetes mellitus |
| | Blindness due to type 1 diabetes mellitus |
| | Lesion of skin due to diabetes mellitus |
| | Pre-existing type 1 diabetes mellitus |
| | Neuropathic arthropathy due to diabetes mellitus |

| | |
|---|---|
| | Chronic kidney disease stage 2 due to type 2 diabetes mellitus |
| | Peripheral neuropathy due to type 2 diabetes mellitus |
| | Moderate nonproliferative retinopathy due to secondary diabetes mellitus |
| | Gangrene due to diabetes mellitus |
| | Hyperosmolar non-ketotic state due to type 2 diabetes mellitus |
| | Severe nonproliferative retinopathy of left eye due to diabetes mellitus |
| | Ulcer of heel due to type 2 diabetes mellitus |
| | Severe nonproliferative retinopathy of right eye due to diabetes mellitus |
| | Chronic kidney disease stage 4 due to type 2 diabetes mellitus |
| | Hyperlipidemia due to type 2 diabetes mellitus |
| | Postpartum gestational diabetes mellitus |
| | Vitreous hemorrhage due to diabetes mellitus |
| | Vitreous hemorrhage of left eye due to diabetes mellitus |
| | Mixed hyperlipidemia due to type 1 diabetes mellitus |
| | Diabetes mellitus in mother complicating childbirth |
| | Chronic kidney disease stage 4 due to type 1 diabetes mellitus |
| | Pre-existing type 2 diabetes mellitus |
| | Erectile dysfunction due to type 2 diabetes mellitus |
| | Mild nonproliferative retinopathy due to secondary diabetes mellitus |
| | Macroalbuminuric nephropathy due to diabetes mellitus |
| | Hypoglycemic coma due to diabetes mellitus |
| | Traction detachment of retina due to type 1 diabetes mellitus |
| | Microalbuminuria due to type 2 diabetes mellitus |
| | Lumbosacral radiculoplexus neuropathy due to type 1 diabetes mellitus |
| | Hyperglycemia due to diabetes mellitus |
| | Disorder of soft tissue due to diabetes mellitus |
| | Nonproliferative retinopathy of left eye due to diabetes mellitus |
| | Ulcer of right foot due to type 2 diabetes mellitus |
| | Abnormal metabolic state due to diabetes mellitus |
| | Hyperglycemic crisis due to diabetes mellitus |
| | Macular edema of left eye due to diabetes mellitus |
| | O/E - right eye stable treated proliferative diabetic retinopathy |
| | Peripheral angiopathy due to type 1 diabetes mellitus |
| | Proteinuria due to type 2 diabetes mellitus |
| | Ulcer of left foot due to type 2 diabetes mellitus |
| | Microalbuminuric diabetic nephropathy |
| | Chronic kidney disease stage 1 due to type 2 diabetes mellitus |
| | Ulcer of lower limb due to type 2 diabetes mellitus |
| | Diabetes mellitus associated with cystic fibrosis |

| | |
|---|---|
| | O/E - left eye stable treated proliferative diabetic retinopathy |
| | Cellulitis of foot due to diabetes mellitus |
| | Type 1 diabetes mellitus with arthropathy |
| | Glomerulopathy due to diabetes mellitus |
| | Hyperosmolar hyperglycemic coma due to diabetes mellitus without ketoacidosis |
| | Chronic painful neuropathy due to diabetes mellitus |
| | Diarrhea due to diabetes mellitus |
| | Skin ulcer of toe due to diabetes mellitus type 1 |
| | Hyperosmolar coma due to type 1 diabetes mellitus |
| | Diabetes mellitus associated with hormonal etiology |
| | Retinopathy due to secondary diabetes mellitus |
| | Dermatitis due to drug induced diabetes mellitus |
| | Ulcer of foot due to type 1 diabetes mellitus |
| | Insulin dependent diabetes mellitus type 1A |
| | Acidosis due to type 2 diabetes mellitus |
| | Diabetes mellitus induced by non-steroid drugs |
| | Chronic kidney disease stage 5 due to type 2 diabetes mellitus |
| | Bullosis diabeticorum |
| | Hyperosmolar non-ketotic state due to diabetes mellitus |
| | Peripheral sensory neuropathy due to type 2 diabetes mellitus |
| | Skin ulcer due to type 2 diabetes mellitus |
| | Nephrotic syndrome due to diabetes mellitus |
| | Maturity-onset diabetes of the young, type 5 |
| | Diabetes mellitus due to cystic fibrosis |
| | Microalbuminuria due to type 1 diabetes mellitus |
| | Diabetes mellitus in the puerperium - baby delivered during previous episode of care |
| | Diabetes mellitus associated with pancreatic disease |
| | Hypertension in chronic kidney disease stage 3 due to type 2 diabetes mellitus |
| | Hypoglycemic event due to diabetes |
| | O/E - left eye background diabetic retinopathy |
| | Coronary artery disease due to type 2 diabetes mellitus |
| | Glaucoma due to type 2 diabetes mellitus |
| | **Diagnostic concept name** |
| **Hypertension (HTN)** | |
| | Essential hypertension |

| | |
|---|---|
| | Hypertensive heart failure |
| | Hypertensive heart and renal disease with (congestive) heart failure |
| | Benign essential hypertension |
| | Hypertensive urgency |
| | Hypertensive disorder |
| | Hypertensive heart disease without congestive heart failure |
| | Hypertensive emergency |
| | Hypertensive heart disease with congestive heart failure |
| | Hypertensive heart and chronic kidney disease |
| | Hypertensive renal disease |
| | Hypertensive heart AND renal disease |
| | Hypertensive retinopathy |
| | Benign hypertensive renal disease with renal failure |
| | Hypertensive heart disease |
| | Benign hypertension |
| | Hypertensive encephalopathy |
| | Hypertensive renal failure |
| | Malignant essential hypertension |
| | Hypertensive crisis |
| | Hypertensive heart AND chronic kidney disease with congestive heart failure |
| | Hypertensive heart and renal disease with renal failure |
| | Benign hypertensive heart disease without congestive heart failure |
| | Benign hypertensive renal disease |
| | Benign hypertensive heart AND renal disease |
| | Hypertensive heart AND chronic kidney disease stage 5 |
| | Malignant hypertension |
| | Benign hypertensive heart disease with congestive cardiac failure |
| | Blind hypertensive eye |
| | Hypertensive heart AND chronic kidney disease stage 3 |
| | Pre-existing hypertensive chronic kidney disease in mother complicating pregnancy |
| | Malignant hypertensive heart disease without congestive heart failure |
| | Pre-existing hypertensive heart disease complicating pregnancy, childbirth and the puerperium |
| | Hypertensive left ventricular hypertrophy |
| | Hypertensive complication |
| | Hypertensive heart AND chronic kidney disease stage 4 |
| | Malignant hypertensive heart AND renal disease |
| | Pre-existing hypertensive heart disease in mother complicating pregnancy |
| | Labile essential hypertension |

| | |
|---|---|
| | Benign hypertensive heart disease |
| | Resistant hypertensive disorder |
| | Pre-existing hypertensive heart and chronic kidney disease in mother complicating childbirth |
| | Hypertensive heart AND chronic kidney disease stage 2 |
| | Pre-existing hypertensive heart and chronic kidney disease in mother complicating pregnancy |
| | Hypertensive nephrosclerosis |
| | Hypertensive heart and renal disease with both (congestive) heart failure and renal failure |
| | Pre-existing hypertensive heart and renal disease complicating pregnancy, childbirth and the puerperium |
| | Malignant hypertensive heart disease |
| | **Diagnostic concept name** |
| **Cardiovascular disease (CVD)** | |
| | Congestive heart failure |
| | Atherosclerosis of coronary artery without angina pectoris |
| | Atrial fibrillation |
| | Paroxysmal atrial fibrillation |
| | Heart failure |
| | Chronic systolic heart failure |
| | Chronic diastolic heart failure |
| | Chronic congestive heart failure |
| | Hypertensive heart failure |
| | Chronic atrial fibrillation |
| | Cardiomyopathy |
| | Hypertensive heart and renal disease with (congestive) heart failure |
| | Old myocardial infarction |
| | Generalized ischemic myocardial dysfunction |
| | Acute on chronic diastolic heart failure |
| | Cardiomegaly |
| | Acute on chronic systolic heart failure |
| | Non-rheumatic aortic sclerosis |
| | Chronic combined systolic and diastolic heart failure |
| | Cardiac arrhythmia |
| | Diastolic heart failure |
| | Persistent atrial fibrillation |
| | Acute non-ST segment elevation myocardial infarction |

| | |
|---|---|
| | Ventricular tachycardia |
| | Atrial flutter |
| | Non-rheumatic mitral valve stenosis with regurgitation |
| | Angina co-occurrent and due to coronary arteriosclerosis |
| | Supraventricular tachycardia |
| | Sick sinus syndrome |
| | Paralytic syndrome on one side of the body as late effect of cerebrovascular accident |
| | Acute on chronic combined systolic and diastolic heart failure |
| | Coronary arteriosclerosis |
| | Dilated cardiomyopathy |
| | Angina pectoris |
| | Systolic heart failure |
| | Ventricular premature complex |
| | Heart disease |
| | Right bundle branch block |
| | Coronary atherosclerosis |
| | Complete atrioventricular block |
| | Left bundle branch block |
| | First degree atrioventricular block |
| | Arteriosclerosis of coronary artery bypass graft |
| | Aortic incompetence, non-rheumatic |
| | Acute systolic heart failure |
| | Acute ST segment elevation myocardial infarction |
| | Tricuspid incompetence, non-rheumatic |
| | Acute ischemic heart disease |
| | Acute diastolic heart failure |
| | Cardiac transplant disorder |
| | Permanent atrial fibrillation |
| | Chronic ischemic heart disease |
| | Long QT syndrome |
| | Unstable angina co-occurrent and due to coronary arteriosclerosis |
| | Atrial premature complex |
| | Hypertensive heart disease without congestive heart failure |
| | Non-rheumatic heart valve disorder |
| | Rheumatic tricuspid valve regurgitation |
| | Cardiomyopathy associated with another disorder |
| | Typical atrial flutter |
| | Rheumatic disease of heart valve |

| | |
|---|---|
| | Late effects of cerebrovascular disease |
| | Preinfarction syndrome |
| | Second degree atrioventricular block |
| | Residual cognitive deficit as late effect of cerebrovascular accident |
| | Cardiac transplant rejection |
| | Mural thrombus of heart |
| | Hypertrophic cardiomyopathy |
| | Myocardial infarction due to demand ischemia |
| | Atrial septal defect |
| | Disorders of both mitral and tricuspid valves |
| | Aphasia as late effect of cerebrovascular disease |
| | Cardiac arrest |
| | Right ventricular failure |
| | Non-rheumatic mitral valve disease |
| | Atypical atrial flutter |
| | Conduction disorder of the heart |
| | Biventricular congestive heart failure |
| | Hypertensive heart disease with congestive heart failure |
| | Rheumatic disease of mitral AND aortic valves |
| | Valvular endocarditis |
| | Atrioventricular block |
| | Primary cardiomyopathy |
| | Dysphagia as a late effect of cerebrovascular accident |
| | Acute exacerbation of chronic congestive heart failure |
| | Hypertrophic obstructive cardiomyopathy |
| | Rheumatic disease of mitral valve |
| | Left anterior fascicular block |
| | Left heart failure |
| | Hypertensive heart and chronic kidney disease |
| | Bifascicular block |
| | Acute combined systolic and diastolic heart failure |
| | Combined disorders of mitral, aortic and tricuspid valves |
| | Ventricular fibrillation |
| | Chronic pulmonary heart disease |
| | Coronary arteriosclerosis in artery of transplanted heart |
| | Longstanding persistent atrial fibrillation |
| | Aortic valve disorder |
| | Acute myocardial infarction |
| | Congenital heart disease |

| | |
|---|---|
| | Paroxysmal ventricular tachycardia |
| | Infective endocarditis |
| | Myocardial infarction |
| | Acute cor pulmonale |
| | Mitral valve prolapse |
| | Hypertensive heart AND renal disease |
| | Takotsubo cardiomyopathy |
| | Congenital insufficiency of aortic valve |
| | Acute subendocardial infarction |
| | Dysarthria as late effects of cerebrovascular disease |
| | Mitral valve disorder |
| | Weakness of face muscles as sequela of stroke |
| | Chronic right-sided heart failure |
| | Sinus node dysfunction |
| | Rheumatic mitral stenosis |
| | Chronic total occlusion of coronary artery |
| | Pulmonary incompetence, non-rheumatic |
| | Non-rheumatic mitral regurgitation |
| | Aortic stenosis, non-rheumatic |
| | Acute on chronic right-sided congestive heart failure |
| | Chronic cor pulmonale |
| | Tetralogy of Fallot |
| | Acute ST segment elevation myocardial infarction involving left anterior descending coronary artery |
| | Hypertensive heart disease |
| | Ischemic myocardial dysfunction |
| | Intraventricular conduction defect |
| | Non-rheumatic mitral valve stenosis |
| | Sarcoid heart muscle disease |
| | Rheumatic heart disease |
| | Aortic valve stenosis |
| | Acute ST segment elevation myocardial infarction due to right coronary artery occlusion |
| | Congenital anomaly of coronary artery |
| | Ventricular tachyarrhythmia |
| | Coronary artery spasm |
| | Acute congestive heart failure |
| | Saddle embolus of pulmonary artery with acute cor pulmonale |
| | Heart failure with normal ejection fraction |
| | Multiple valve disease |

| | |
|---|---|
| | Cardiac transplant failure |
| | Heart block |
| | Ventricular septal defect |
| | Aortic valve stenosis with insufficiency |
| | Speech and language deficit as late effect of cerebrovascular accident |
| | Atrial fibrillation with rapid ventricular response |
| | Cardiovascular symptoms |
| | Disorders of both aortic and tricuspid valves |
| | Paroxysmal supraventricular tachycardia |
| | Arteriosclerosis of autologous vein coronary artery bypass graft |
| | Aortic valve regurgitation |
| | Restrictive cardiomyopathy |
| | Dilated cardiomyopathy secondary to peripartum heart disease |
| | Ataxia as sequela of cerebrovascular disease |
| | Dilated cardiomyopathy secondary to alcohol |
| | Acute right-sided heart failure |
| | Coronary thrombosis not resulting in myocardial infarction |
| | Pulmonary heart disease |
| | Dissection of coronary artery |
| | Premature beats |
| | Ventricular premature beats |
| | Discordant ventriculoarterial connection |
| | Mitral valve regurgitation |
| | Congestive heart failure due to left ventricular systolic dysfunction |
| | Symptomatic congestive heart failure |
| | Aneurysm of heart |
| | Cardiac tamponade |
| | Ebstein's anomaly |
| | Right heart failure secondary to left heart failure |
| | Aberrant premature complexes |
| | Accelerated atrioventricular conduction |
| | Paroxysmal tachycardia |
| | Myocarditis |
| | Chronic heart failure |
| | Disorder of transplanted heart |
| | Rheumatic mitral regurgitation |
| | Atrioventricular septal defect and common atrioventricular junction |
| | Rheumatic disease of tricuspid valve |
| | Disorder of coronary artery |

| | |
|---|---|
| | Hemiplegia of nondominant side as late effect of cerebrovascular disease |
| | Acute and subacute endocarditis |
| | Rheumatic mitral stenosis with regurgitation |
| | Chronic Chagas disease with heart involvement |
| | Postoperative cardiac complication |
| | Thrombosis of atrium, auricular appendage, and ventricle due to and following acute myocardial infarction |
| | Stable angina |
| | Acute heart failure |
| | Nonsustained ventricular tachycardia |
| | Cardiomyopathy due to viral infection |
| | Hemiplegia as late effect of cerebrovascular disease |
| | Tricuspid valve disorder, non-rheumatic |
| | Congenital stenosis of tricuspid valve |
| | Exacerbation of congestive heart failure |
| | Hypertensive heart AND chronic kidney disease with congestive heart failure |
| | Dextrocardia |
| | Cardiac arrest due to cardiac disorder |
| | Acute coronary syndrome |
| | Cardiovascular stress test abnormal |
| | Ostium secundum type atrial septal defect |
| | Common arterial trunk (truncus arteriosus) |
| | Left ventricular thrombus |
| | Pulmonary stenosis, non-rheumatic |
| | Mitral valve stenosis |
| | Chronic heart failure co-occurrent with normal ejection fraction |
| | Hemiplegia of dominant side as late effect of cerebrovascular disease |
| | Trifascicular block |
| | Decompensated cardiac failure |
| | Subsequent non-ST segment elevation myocardial infarction |
| | Rheumatic aortic stenosis |
| | Arteriosclerosis of autologous arterial coronary artery bypass graft |
| | Myocarditis due to infectious agent |
| | Cardiovascular sequelae of disorders |
| | Hemiparesis as late effect of cerebrovascular accident |
| | Left posterior fascicular block |
| | Aphasia as late effect of cerebrovascular accident |
| | Dysphasia as late effect of cerebrovascular disease |
| | Acute and subacute bacterial endocarditis |

| | |
|---|---|
| | Heart failure with reduced ejection fraction |
| | Recurrent coronary arteriosclerosis after percutaneous transluminal coronary angioplasty |
| | Monoplegia of nondominant upper limb as a late effect of cerebrovascular accident |
| | Double inlet ventricle |
| | Calcification of coronary artery |
| | Left ventricular hypertrophy |
| | Congenital stenosis of aortic valve |
| | Eisenmenger's syndrome |
| | Tricuspid valve disorder |
| | Endocarditis |
| | Postcardiotomy syndrome |
| | Left ventricular cardiac dysfunction |
| | Disorder of prosthetic cardiac valve |
| | Acute ST segment elevation myocardial infarction due to left coronary artery occlusion |
| | Congenital subaortic stenosis |
| | Hypertensive heart and renal disease with renal failure |
| | Benign hypertensive heart disease without congestive heart failure |
| | Acute on chronic heart failure co-occurrent with normal ejection fraction |
| | Heart valve disorder |
| | Ischemic heart disease |
| | Congenital anomaly of heart valve |
| | Bicuspid aortic valve |
| | Monoplegia of dominant upper limb as a late effect of cerebrovascular accident |
| | Non-rheumatic mitral valve prolapse |
| | Mobitz type II atrioventricular block |
| | Thallium stress test abnormal |
| | Pulmonary valve disorder |
| | Aneurysm of coronary vessels |
| | Atrial tachycardia |
| | Congenital stenosis of mitral valve |
| | Hemiplegia as late effect of cerebrovascular accident |
| | Monoplegia of dominant lower limb as a late effect of cerebrovascular accident |
| | Benign neoplasm of heart |
| | Prosthetic cardiac paravalvular leak |
| | Atresia of pulmonary valve |
| | Rapid atrial fibrillation |

| | |
|---|---|
| | Tricuspid valve regurgitation |
| | Cardiac sarcoidosis |
| | High output heart failure |
| | Atrial fibrillation and flutter |
| | Congenital anomaly of tricuspid valve |
| | Dysfunction of right cardiac ventricle |
| | Arteriosclerosis of coronary artery bypass graft of transplanted heart |
| | Mural thrombus of left ventricle |
| | Left ventricular systolic dysfunction |
| | Supraventricular premature beats |
| | Acute myocardial infarction of anterior wall |
| | Rheumatic endocarditis |
| | Mechanical complication of heart valve prosthesis |
| | Benign hypertensive heart AND renal disease |
| | Contusion to heart |
| | Injury of heart |
| | Hypertensive heart AND chronic kidney disease stage 5 |
| | Junctional premature complex |
| | Monoplegia of nondominant lower limb as a late effect of cerebrovascular accident |
| | Tricuspid stenosis, non-rheumatic |
| | Acute rheumatic endocarditis |
| | Double outlet right ventricle |
| | Calcific coronary arteriosclerosis |
| | Thrombus of left atrium |
| | Chagas' disease with heart involvement |
| | Bundle branch block |
| | Cardiac insufficiency following cardiac surgery |
| | Paralytic syndrome as late effect of stroke |
| | Patent foramen ovale |
| | Left ventricular myocardial noncompaction cardiomyopathy |
| | Acute myocarditis |
| | Abnormality of fetal heart |
| | Abscess of aortic valve |
| | Arteriosclerosis of arterial coronary artery bypass graft |
| | Rheumatic disease of aortic valve |
| | Sinus bradycardia |
| | Premature atrial contraction |
| | Left bundle branch hemiblock |

| | |
|---|---|
| | Sudden cardiac death |
| | Decompensated chronic heart failure |
| | Congenital malposition of heart |
| | Aortic valve sclerosis |
| | Coronary graft stenosis |
| | Ostium primum defect |
| | Hypoplastic left heart syndrome |
| | Nutritional and metabolic cardiomyopathies |
| | Acute bacterial endocarditis |
| | Acquired cardiac septal defect |
| | Slow ventricular response |
| | Congenital atresia of pulmonary valve |
| | Acute myocardial infarction of inferior wall |
| | Atypical angina |
| | Benign hypertensive heart disease with congestive cardiac failure |
| | Sequela of cerebrovascular accident |
| | Injury of heart with hemopericardium |
| | Cleft leaflet of mitral valve |
| | Primary eosinophilic endomyocardial restrictive cardiomyopathy |
| | Candidal endocarditis |
| | Left ventricular diastolic dysfunction |
| | Subaortic stenosis |
| | Weakness as a late effect of stroke |
| | Tachycardia-bradycardia |
| | Angina co-occurrent and due to arteriosclerosis of coronary artery bypass graft |
| | Lipid-rich atherosclerosis of coronary artery |
| | Rheumatic aortic regurgitation |
| | Cardiac arrest during surgery |
| | Paroxysmal atrial flutter |
| | Atrial septal defect due to and following acute myocardial infarction |
| | Primary malignant neoplasm of heart |
| | Acute rejection of cardiac transplant |
| | Acute myocardial infarction of inferoposterior wall |
| | Heart transplant failure and rejection |
| | Ventricular flutter |
| | Subsequent ST segment elevation myocardial infarction |
| | Primary dilated cardiomyopathy |
| | Postpartum cardiomyopathy |

| | |
|---|---|
| | Discordant atrioventricular connection |
| | Hypertensive heart AND chronic kidney disease stage 3 |
| | Congenital heart block |
| | Right bundle branch block AND left anterior fascicular block |
| | Exercise-induced angina |
| | Isolated (Fiedler's) myocarditis |
| | Endocardial fibroelastosis |
| | Congenital stenosis of pulmonary valve |
| | Malignant hypertensive heart disease without congestive heart failure |
| | Pre-existing hypertensive heart disease complicating pregnancy, childbirth and the puerperium |
| | Ventricular bigeminy |
| | Secondary nonischemic congestive cardiomyopathy |
| | Incomplete right bundle branch block |
| | Nonischemic congestive cardiomyopathy |
| | Endocarditis associated with another disorder |
| | Congenital insufficiency of mitral valve |
| | Wolff-Parkinson-White pattern |
| | Disorder of cardiac function |
| | Familial cardiomyopathy |
| | Hypertensive left ventricular hypertrophy |
| | Bilateral bundle branch block |
| | Left atrial enlargement |
| | Right cardiac ventricular dilatation |
| | Congenital septal defect of heart |
| | Atrial paroxysmal tachycardia |
| | Myocardial degeneration |
| | Hypertensive heart AND chronic kidney disease stage 4 |
| | Cardiac complication of procedure |
| | Fetal heart disorder |
| | Monoplegia of lower limb as late effect of cerebrovascular disease |
| | Atrial arrhythmia |
| | Mitral stenosis with insufficiency |
| | Congenital cardiovascular disorder during pregnancy - baby not yet delivered |
| | Re-entry ventricular arrhythmia |
| | Pulmonic valve stenosis |
| | Post cardiac operation functional disturbance |
| | Viral myocarditis |
| | Torsades de pointes |

| | |
|---|---|
| | Malignant hypertensive heart AND renal disease |
| | Fetal dysrhythmia |
| | Rupture of chordae tendineae |
| | Mobitz type I incomplete atrioventricular block |
| | Coronary sinus abnormality |
| | Subsequent myocardial infarction of inferior wall |
| | Hypertrophic cardiomyopathy without obstruction |
| | Mechanical breakdown of prosthetic heart valve |
| | Atrial septal defect through coronary sinus orifice |
| | Senile cardiac amyloidosis |
| | Arteriosclerosis of nonautologous coronary artery bypass graft |
| | Ventricular arrhythmia |
| | Endocarditis due to systemic lupus erythematosus |
| | Acute myocardial infarction of inferolateral wall |
| | Mechanical complication due to heart valve prosthesis |
| | Sequelae of cardiovascular disorders |
| | Pre-existing hypertensive heart disease in mother complicating pregnancy |
| | Right ventricular hypertension |
| | Left ventricular outflow tract obstruction |
| | Wide QRS ventricular tachycardia |
| | Subsequent myocardial infarction of anterior wall |
| | Thrombus of cardiac chamber |
| | Carditis due to rheumatic fever |
| | Refractory heart failure |
| | Multi vessel coronary artery disease |
| | Congenital insufficiency of pulmonary valve |
| | Acute coronary artery occlusion not resulting in myocardial infarction |
| | Seizure disorder as sequela of stroke |
| | Mitral and aortic incompetence |
| | Subacute periendocarditis |
| | Angina, class I |
| | AV nodal re-entry tachycardia |
| | Congestive heart failure stage D |
| | Non-specific intraventricular conduction delay |
| | Disorder of right cardiac ventricle |
| | Sinus arrest |
| | AV-junctional (nodal) bradycardia |
| | Acute myocardial infarction of anterolateral wall |
| | AV junctional rhythm |

| | |
|---|---|
| | Post-infarction ventricular septal defect |
| | Benign hypertensive heart disease |
| | Primary hypertrophic cardiomyopathy |
| | Significant coronary bypass graft disease |
| | Nodular calcific aortic valve stenosis |
| | Fluency disorder as sequela of cerebrovascular disease |
| | Cor pulmonale |
| | Bacterial endocarditis |
| | Williams syndrome |
| | Vegetation of heart |
| | Post infarct angina |
| | Old inferior myocardial infarction |
| | Cardiac arrest as a complication of care |
| | Persistent sinus bradycardia |
| | Brugada syndrome |
| | Abnormality of left atrial appendage |
| | Myocardial ischemia |
| | Prinzmetal angina |
| | Abnormal vision as a late effect of cerebrovascular disease |
| | Heart-lung transplant failure and rejection |
| | Anomalous atrioventricular excitation |
| | Silent myocardial ischemia |
| | Primary endocardial fibroelastosis |
| | Visual disturbance as sequela of cerebrovascular disease |
| | Symptomatic sinus bradycardia |
| | Mitral valve vegetations |
| | Re-entrant atrioventricular node tachycardia |
| | Systolic heart failure stage B |
| | Cognitive deficit due to and following cerebrovascular disease |
| | Re-entrant atrioventricular tachycardia |
| | Papillary fibroelastoma of heart |
| | Papillary fibroelastoma |
| | Staphylococcal endocarditis |
| | Transthyretin related familial amyloid cardiomyopathy |
| | Pulmonic valve regurgitation |
| | Left ventricular aneurysm |
| | Myocardial bridge of coronary artery |
| | Rheumatic aortic stenosis with regurgitation |
| | Mural thrombus of left ventricle following acute myocardial infarction |

| | |
|---|---|
| | Persistent ostium secundum |
| | Atrial septal aneurysm |
| | Pre-existing hypertensive heart and chronic kidney disease in mother complicating childbirth |
| | Severe aortic valve stenosis |
| | Hemiparesis as late effect of cerebrovascular disease |
| | Post-phlebitic dermatosis of lower leg |
| | Rheumatic tricuspid valve stenosis |
| | Non-rheumatic pulmonary valve stenosis with regurgitation |
| | Acute rheumatic pericarditis |
| | Monoplegia of upper limb as late effect of cerebrovascular disease |
| | Congenital pulmonary valve abnormality |
| | Post-infarction mural thrombus |
| | Prosthetic valve endocarditis |
| | Cardiac volume overload |
| | Acute endocarditis |
| | Cardiac complication |
| | Severe sinus bradycardia |
| | Mitral insufficiency and aortic stenosis |
| | Coronary artery bypass graft occlusion |
| | Common ventricle |
| | Prosthetic cardiac valve displacement |
| | Idiopathic myocarditis |
| | Neonatal cardiac failure |
| | Congestive heart failure stage C |
| | Complete atrioventricular block as complication of atrioventricular nodal ablation |
| | Ectopic beats |
| | Sensory disorder as a late effect of cerebrovascular disease |
| | Hypertensive heart AND chronic kidney disease stage 2 |
| | Ectopic atrial beats |
| | Spasticity as sequela of stroke |
| | Severe tricuspid valve regurgitation |
| | Non-rheumatic pulmonary valve disorder |
| | Myxoid transformation of mitral valve |
| | Neonatal tachycardia |
| | Rheumatic tricuspid stenosis and insufficiency |
| | Cardiac insufficiency during AND/OR resulting from a procedure |
| | Ataxia as sequela of cerebrovascular accident |
| | Right atrial dilatation |

| | |
|---|---|
| | Viral endocarditis |
| | Supraventricular arrhythmia |
| | Idiopathic hypertrophic subaortic stenosis |
| | Myxedema heart disease |
| | Angina decubitus |
| | Infundibular pulmonic stenosis |
| | Tachyarrhythmia |
| | Endocardial cushion defect |
| | Ventricular tachycardia with normal heart |
| | Eosinophilic myocarditis |
| | Masses on mitral apparatus |
| | Coronary artery fistula |
| | Valvular cardiomyopathy |
| | Acute Chagas' disease with heart involvement |
| | Prosthetic cardiac valve thrombosis |
| | Moderate aortic valve stenosis |
| | Acute heart failure co-occurrent with normal ejection fraction |
| | Atrioventricular dissociation |
| | Triple vessel disease of the heart |
| | Kyphoscoliotic heart disease |
| | Right hypoplastic heart syndrome |
| | Acute rheumatic heart disease |
| | Acute rheumatic myocarditis |
| | Coronary artery stent thrombosis |
| | Neonatal bradycardia |
| | Chronic right-sided congestive heart failure |
| | Ischemic congestive cardiomyopathy |
| | Pre-existing hypertensive heart and chronic kidney disease in mother complicating pregnancy |
| | Right atrial enlargement |
| | Mild aortic valve regurgitation |
| | Complete transposition of great vessels |
| | Arrhythmogenic right ventricular dysplasia |
| | Myocardial disease |
| | Progressive angina |
| | Typical angina |
| | Aortic valve calcification |
| | Congenital subaortic stenosis due to fibromuscular shelf |
| | Atrial hypertrophy |

| | |
|---|---|
| | Atrial thrombosis |
| | Cardiorenal syndrome |
| | Angina, class II |
| | Cardiac septal defects |
| | Injury of heart without open wound into thorax |
| | Heart disease in mother complicating pregnancy, childbirth AND/OR puerperium |
| | Right bundle branch block AND left posterior fascicular block |
| | Cor triatriatum |
| | Tachycardia-induced cardiomyopathy |
| | Disorder of cardiac ventricle |
| | Cardiac ventricular dilatation |
| | Cardiac disease in pregnancy |
| | Moderate left ventricular systolic dysfunction |
| | Controlled atrial fibrillation |
| | Prosthetic cardiac valve calcification |
| | Subacute endocarditis |
| | Moderate laceration of heart with hemopericardium |
| | Paralytic syndrome of nondominant side as late effect of stroke |
| | Holt-Oram syndrome |
| | Rheumatic myocarditis |
| | Unifocal PVCs |
| | Vertigo as sequela of cerebrovascular disease |
| | Coronary arteriosclerosis after percutaneous coronary angioplasty |
| | Acute right-sided congestive heart failure |
| | Thrombus of right atrium |
| | Myocardial dysfunction |
| | Primary idiopathic dilated cardiomyopathy |
| | Vertigo as late effect of stroke |
| | Paralytic syndrome of dominant side as late effect of stroke |
| | Atrial bigeminy |
| | Isomerism of atrial appendages |
| | Right ventricular diastolic dysfunction |
| | Severe mitral valve regurgitation |
| | Stokes-Adams syndrome |
| | Mitral valve prolapse syndrome |
| | Incomplete left bundle branch block |
| | Non-rheumatic tricuspid valve stenosis with insufficiency |
| | Common atrium |

| | |
|---|---|
| | Chronic bacterial endocarditis |
| | Severe mitral valve stenosis |
| | Moderate mitral valve regurgitation |
| | Hypertrophic cardiomegaly |
| | Acute myocardial infarction of inferior wall involving right ventricle |
| | Hyperkinetic heart disease |
| | Congenital cardiovascular disorders during pregnancy, childbirth and the puerperium |
| | Mitral and aortic stenosis |
| | Non-obstructive atherosclerosis of coronary artery |
| | Tuberculosis of heart |
| | Bulbus cordis and cardiac septal closure anomalies |
| | Prosthetic aortic valve regurgitation |
| | Sustained ventricular tachycardia |
| | Isolated diffuse granulomatous myocarditis |
| | New onset angina |
| | Cardiac arrest during AND/OR resulting from a procedure |
| | Hypertensive heart and renal disease with both (congestive) heart failure and renal failure |
| | Left main coronary artery disease |
| | Neurogenic bladder as late effect of cerebrovascular accident |
| | Atrial myxoma |
| | Acute ST segment elevation myocardial infarction due to occlusion of circumflex coronary artery |
| | Asystole |
| | Complete left bundle branch block |
| | Tricuspid valve vegetations |
| | D - transposition of the great vessels |
| | Congestive heart failure with right heart failure |
| | Mitral valve annular calcification |
| | Acute ST segment elevation myocardial infarction of inferior wall |
| | Acute left-sided congestive heart failure |
| | Myxoid transformation of cardiac valve |
| | Mild tricuspid valve regurgitation |
| | Myocarditis due to influenza virus |
| | Malignant hypertensive heart disease |
| | Atrial dilatation |
| | L - transposition of the great vessels |
| | Mixed myocardial ischemia and infarction |
| | Dysarthria due to and following cerebrovascular accident |

| | |
|---|---|
| | Bilateral enlargement of atria |
| | Ventricular tachycardia, polymorphic |
| | Acute myocarditis associated with another disorder |
| | Heart valve regurgitation |
| | Infection of cardiac graft |
| | Pre-existing hypertensive heart and renal disease complicating pregnancy, childbirth and the puerperium |
| | Coronary artery disease due to type 2 diabetes mellitus |
| | Low output heart failure |
| | Ventricular tachycardia, monomorphic |
| | Mild mitral valve regurgitation |
| | Syphilitic endocarditis |
| | Severe aortic valve regurgitation |
| | Right coronary artery occlusion |
| | Nonsustained paroxysmal ventricular tachycardia |
| | Laceration of heart |
| | Coronary arteriosclerosis following coronary artery bypass graft |
| | Nodal rhythm disorder |
| | Incomplete atrioventricular block with atrioventricular response |
| | Electromechanical dissociation |
| | Subacute bacterial endocarditis |
| | **Diagnostic concept name** |
| **Heart Failure (HF)** | |
| | Congestive heart failure |
| | Heart failure |
| | Chronic systolic heart failure |
| | Chronic diastolic heart failure |
| | Chronic congestive heart failure |
| | Hypertensive heart failure |
| | Hypertensive heart and renal disease with (congestive) heart failure |
| | Acute on chronic diastolic heart failure |
| | Acute on chronic systolic heart failure |
| | Chronic combined systolic and diastolic heart failure |
| | Diastolic heart failure |
| | Acute on chronic combined systolic and diastolic heart failure |
| | Systolic heart failure |
| | Acute systolic heart failure |
| | Acute diastolic heart failure |

| | |
|---|---|
| | Hypertensive heart disease without congestive heart failure |
| | Right ventricular failure |
| | Biventricular congestive heart failure |
| | Hypertensive heart disease with congestive heart failure |
| | Acute exacerbation of chronic congestive heart failure |
| | Left heart failure |
| | Acute combined systolic and diastolic heart failure |
| | Acute cor pulmonale |
| | Chronic right-sided heart failure |
| | Acute on chronic right-sided congestive heart failure |
| | Chronic cor pulmonale |
| | Acute congestive heart failure |
| | Saddle embolus of pulmonary artery with acute cor pulmonale |
| | Heart failure with normal ejection fraction |
| | Acute right-sided heart failure |
| | Congestive heart failure due to left ventricular systolic dysfunction |
| | Symptomatic congestive heart failure |
| | Right heart failure secondary to left heart failure |
| | Chronic heart failure |
| | Acute heart failure |
| | Exacerbation of congestive heart failure |
| | Hypertensive heart AND chronic kidney disease with congestive heart failure |
| | Chronic heart failure co-occurrent with normal ejection fraction |
| | Decompensated cardiac failure |
| | Heart failure with reduced ejection fraction |
| | Benign hypertensive heart disease without congestive heart failure |
| | Acute on chronic heart failure co-occurrent with normal ejection fraction |
| | High output heart failure |
| | Cardiac insufficiency following cardiac surgery |
| | Decompensated chronic heart failure |
| | Benign hypertensive heart disease with congestive cardiac failure |
| | Malignant hypertensive heart disease without congestive heart failure |
| | Refractory heart failure |
| | Congestive heart failure stage D |
| | Cor pulmonale |
| | Systolic heart failure stage B |
| | Congestive heart failure stage C |
| | Neonatal cardiac failure |
| | Cardiac insufficiency during AND/OR resulting from a procedure |

| | |
|---|---|
| | Chronic right-sided congestive heart failure |
| | Acute heart failure co-occurrent with normal ejection fraction |
| | Cardiorenal syndrome |
| | Acute right-sided congestive heart failure |
| | Hypertensive heart and renal disease with both (congestive) heart failure and renal failure |
| | Acute left-sided congestive heart failure |
| | Low output heart failure |
| | Congestive heart failure with right heart failure |
| | **Diagnostic concept name** |
| **Sepsis** | |
| | Sepsis caused by Staphylococcus without acute organ dysfunction<br>Sepsis during labor, delivered<br>Septic shock<br>Postprocedural septic shock<br>Intrauterine sepsis of fetus<br>Sepsis caused by Acinetobacter baumannii<br>Septic shock co-occurrent with acute organ dysfunction due to methicillin susceptible Staphylococcus aureus<br>Sepsis<br>Septic shock co-occurrent with acute organ dysfunction due to Haemophilus influenzae<br>Amber flag sepsis<br>Sepsis of newborn due to group B Streptococcus<br>Sepsis due to ectopic pregnancy<br>Sepsis due to Pseudomonas<br>Meningococcemia<br>Streptococcal toxic shock syndrome<br>Septic shock co-occurrent with acute organ dysfunction due to Chromobacterium<br>Late-onset neonatal sepsis<br>Puerperal sepsis with postnatal complication<br>Gonococcal arthritis dermatitis syndrome<br>Sepsis due to disease caused by Severe acute respiratory syndrome coronavirus 2<br>Proteus septicemia<br>Sepsis due to Streptococcus pyogenes<br>Post-splenectomy sepsis<br>Sepsis due to Staphylococcus<br>Septic shock co-occurrent with acute organ dysfunction due to Streptococcus<br>Septic shock co-occurrent with acute organ dysfunction due to Staphylococcus<br>Gas gangrene septicemia<br>Sepsis of newborn due to anaerobes<br>Hyperdynamic septic shock |

| | |
|---|---|
| | Meningococcal meningitis with acute meningococcal septicemia<br>Induced termination of pregnancy complicated by sepsis<br>Septic shock co-occurrent with acute organ dysfunction due to Gonococcus<br>Systemic inflammatory response syndrome<br>Sepsis without septic shock<br>Sepsis of fetus caused by Streptococcus pyogenes<br>Septic shock co-occurrent with acute organ dysfunction due to coagulase-negative Staphylococcus<br>Sepsis during labor with antenatal problem<br>Sepsis without acute organ dysfunction caused by Streptococcus pneumoniae<br>Coagulase negative staphylococcus bacteremia<br>Sepsis without acute organ dysfunction<br>Illegal termination of pregnancy with septic shock<br>Sepsis of newborn due to Staphylococcus aureus<br>Sepsis caused by Klebsiella pneumoniae<br>Uncomplicated sepsis<br>Brazilian purpuric fever<br>Sepsis with cutaneous manifestations<br>Septicemic pasteurellosis<br>Endotoxic shock<br>Sepsis caused by Peptostreptococcus<br>Tracheostomy sepsis<br>Puerperal pelvic sepsis<br>Sepsis following infusion, injection, transfusion AND/OR vaccination<br>Postoperative septic shock<br>Postoperative endotoxic shock<br>Sepsis due to Acinetobacter<br>Toxic shock syndrome<br>Sepsis associated with acquired immunodeficiency syndrome<br>Sepsis due to Serratia<br>Overwhelming infection in asplenic patient<br>Sepsis of the newborn<br>CLABSI - central line associated bloodstream infection<br>Systemic inflammatory response syndrome of non-infectious origin without organ failure<br>Induced termination of pregnancy complicated by septic shock<br>Non-infectious systemic inflammatory response syndrome<br>Sepsis associated with internal vascular access<br>Sepsis due to Haemophilus influenzae type B<br>Catheter related bloodstream infection<br>Sepsis in asplenic subject<br>Campylobacter bacteremia<br>Neonatal sepsis due to Streptococcus |

| | |
|---|---|
| | Septic shock co-occurrent with acute organ dysfunction due to anaerobic bacteria<br>Septicemia associated with vascular access catheter<br>Sepsis following molar AND/OR ectopic pregnancy<br>Sepsis due to Actinomyces<br>Illegal termination of pregnancy with sepsis<br>Dengue shock syndrome<br>Sepsis without acute organ dysfunction caused by Serratia species<br>Septicemia due to Erysipelothrix insidiosa<br>Septicemic plague<br>Neonatal sepsis caused by Malassezia<br>Septicemic melioidosis<br>Septic shock co-occurrent with acute organ dysfunction due to Pneumococcus<br>Sepsis due to Streptococcus pneumoniae<br>Infectious systemic inflammatory response syndrome with organ failure<br>Septic shock co-occurrent with acute organ dysfunction due to Serratia<br>Toxic shock syndrome due to methicillin resistant Staphylococcus aureus infection<br>Bacterial sepsis<br>Septic shock co-occurrent with acute organ dysfunction due to Pseudomonas<br>Systemic inflammatory response syndrome without organ dysfunction<br>Sepsis of neonate caused by Streptococcus pyogenes<br>Pyemia<br>Sepsis due to methicillin resistant Staphylococcus aureus<br>Neonatal sepsis caused by Streptococcus<br>Pyrogenic shock<br>Bacteremia due to Salmonella<br>Septic shock co-occurrent with acute organ dysfunction due to Group B streptococcus<br>Systemic inflammatory response syndrome of non-infectious origin with organ failure<br>Sepsis due to Escherichia coli<br>Sepsis of newborn due to Streptococcus agalactiae<br>Septicemic glanders<br>Bacteremia associated with intravascular line<br>Bacterial sepsis of newborn<br>Failed attempted termination of pregnancy with septic shock<br>Sepsis due to Streptococcus group D<br>Perinatal sepsis caused by Streptococcus agalactiae<br>Septicemia due to Bacteroides<br>Gram positive sepsis<br>Neutropenic sepsis<br>Non-infectious systemic inflammatory response syndrome without acute organ failure<br>Bacteremia caused by Gram-positive bacteria |

| | |
|---|---|
| | Sepsis due to Candida<br>Sepsis due to incomplete miscarriage<br>Legal termination of pregnancy with sepsis<br>Perinatal sepsis<br>Biliary sepsis<br>Sepsis due to Gram negative bacteria<br>Legal termination of pregnancy with septic shock<br>Infectious systemic inflammatory response syndrome without organ failure<br>Line sepsis associated with dialysis catheter<br>Septic shock co-occurrent with acute organ dysfunction due to methicillin resistant Staphylococcus aureus<br>Neonatal sepsis caused by Staphylococcus<br>Miscarriage with sepsis<br>Septic shock following molar AND/OR ectopic pregnancy<br>Sepsis during labor<br>Septic shock co-occurrent with acute organ dysfunction due to Group A streptococcus<br>Sepsis due to Streptococcus<br>Severe sepsis<br>Failed attempted termination of pregnancy with sepsis<br>Red flag sepsis<br>Recurrent salmonella sepsis co-occurrent with human immunodeficiency virus infection<br>Early-onset neonatal sepsis<br>Neonatal sepsis due to Staphylococcus<br>Gonococcemia<br>Meningococcal meningitis with meningococcal septicemia<br>Toxic shock syndrome due to methicillin susceptible Staphylococcus aureus<br>Sepsis due to urinary tract infection<br>Sepsis following obstructed labor<br>Infection of hemodialysis tunneled catheter<br>Septic shock co-occurrent with acute organ dysfunction due to Meningococcus<br>Sepsis due to Streptococcus agalactiae<br>Acute tubulointerstitial nephritis associated with systemic infection<br>Septicemia due to Chromobacterium<br>Sepsis due to methicillin-sensitive Staphylococcus aureus<br>Sepsis due to Erysipelothrix<br>Sepsis due to Haemophilus influenzae<br>Sepsis due to oral infection<br>Sepsis due to Staphylococcus aureus<br>Acute meningococcemia<br>Septic shock co-occurrent with acute organ dysfunction due to Gram-positive coccus |

| | |
|---|---|
| | Sepsis due to Streptococcus suis<br>Staphylococcal toxic shock syndrome<br>Sepsis due to Enterobacter<br>Sepsis due to Salmonella<br>Sepsis due to Listeria monocytogenes<br>Bacteremia due to Staphylococcus aureus<br>Sepsis caused by anaerobic streptococcus<br>Perinatal sepsis caused by Escherichia coli<br>Sepsis of newborn due to Escherichia coli<br>Septic shock due to transfusion<br>Sepsis due to fungus<br>Bacteremia<br>Pseudomonas septicemia with skin involvement<br>Sepsis due to anaerobic bacteria<br>Puerperal sepsis<br>Coliform septicemia<br>Vancomycin resistant enterococcal septicemia<br>Postoperative sepsis<br>Sepsis due to coagulase negative Staphylococcus<br>Recurrent salmonella septicemia<br>Bacteremia due to Methicillin resistant Staphylococcus aureus<br>Septicemia due to enterococcus<br>Hypodynamic septic shock<br>Bacteremia caused by Gram-negative bacteria<br>Septic shock co-occurrent with acute organ dysfunction due to Enterococcus<br>Sepsis due to Bacillus anthracis<br>Transient respiratory distress with sepsis<br>Systemic inflammatory response syndrome associated with organ dysfunction<br>Sepsis due to herpes simplex<br>Sepsis caused by virus<br>Sepsis caused by Pseudomonas aeruginosa |
| | **Diagnostic concept name** |
| **End Stage Renal Disease (ESRD)** | |
| | End-stage renal disease |
| | End stage renal failure on dialysis |
| | End stage renal disease due to hypertension |
| | Chronic kidney disease stage 5 on dialysis |
| | Anemia in end stage renal disease |

| | |
|---|---|
| | Malignant hypertensive end stage renal disease |
| | End stage renal failure with renal transplant |
| | Hypertension concurrent and due to end stage renal disease on dialysis |
| | End stage renal disease on dialysis due to type 2 diabetes mellitus |
| | Hypertensive end stage renal disease |
| | Hypertension concurrent and due to end stage renal disease on dialysis due to type 2 diabetes mellitus |
| | Malignant hypertensive end stage renal disease on dialysis |
| | End stage renal disease on dialysis due to type 1 diabetes mellitus |
| | End stage renal disease due to benign hypertension |
| | **Procedure concept name** |
| **Invasive ventilation (IMV)** | |
| | Ventilation assist and management, initiation of pressure or volume preset ventilators for assisted or controlled breathing; hospital inpatient/observation, each subsequent day |
| | Intubation, endotracheal, emergency procedure |
| | Respiratory Ventilation, Greater than 96 Consecutive Hours |
| | Ventilation assist and management, initiation of pressure or volume preset ventilators for assisted or controlled breathing; hospital inpatient/observation, initial day |
| | Dependence on respirator |
| | Respiratory Ventilation, 24-96 Consecutive Hours |
| | Artificial respiration |
| | Respiratory Ventilation, Less than 24 Consecutive Hours |
| | Ventilator finding |
| | Assistance with Respiratory Ventilation, Less than 24 Consecutive Hours |
| | Provision of mechanical ventilator |
| | Assistance with Respiratory Ventilation, 24-96 Consecutive Hours |
| | Assistance with Respiratory Ventilation, Greater than 96 Consecutive Hours |
| | Dependence on ventilator |
| | Complication of ventilation therapy |
| | Endotracheal tube present |
| | **Procedure concept name** |
| **Renal Replacement Therapy (RRT)** | |

| | |
|---|---|
| | Hemodialysis procedure with single evaluation by a physician or other qualified health care professional |
| | Dialysis procedure other than hemodialysis (eg, peritoneal dialysis, hemofiltration, or other continuous renal replacement therapies), with single evaluation by a physician or other qualified health care professional |
| | Unlisted dialysis procedure, inpatient or outpatient |
| | Dialysis procedure other than hemodialysis (eg, peritoneal dialysis, hemofiltration, or other continuous renal replacement therapies) requiring repeated evaluations by a physician or other qualified health care professional, with or without substantial revision of dialysis prescription |
| | Hemodialysis procedure requiring repeated evaluation(s) with or without substantial revision of dialysis prescription |
| | Unscheduled or emergency dialysis treatment for an esrd patient in a hospital outpatient department that is not certified as an esrd facility |
| | Hemodialysis |
| | Dialysis procedure |
| | Continuous venovenous hemodiafiltration |
| | Automated peritoneal dialysis |
| | Peritoneal dialysis catheter maintenance |
| | Dialysis procedure at a medicare certified esrd facility for acute kidney injury without esrd |
| | Ultrafiltration |
| | Peritoneal dialysis |
| | Hemodialysis, maintenance at home |
| | Hemodialysis access flow study to determine blood flow in grafts and arteriovenous fistulae by an indicator method |
| | Renal dialysis |
| | Continuous cycling peritoneal dialysis |
| | Continuous ambulatory peritoneal dialysis |
| | **Drug concept name** |
| **Vasopressors (VP)** | |
| | phenylephrine |
| | norepinephrine |
| | epinephrine |
| | milrinone |
| | vasopressin (USP) |
| | dobutamine |
| | dopamine |

**Supplementary Table 2. Descriptive characteristics of hospitalized COVID positive patients with and without AKI by the two AKI definitions.**

| **Variables** | **AKI both criteria (N=**50889) | **AKI – Code-based only (N=**20875) | **AKI – SCr-based only (N=**54714) | **No AKI (by either criterion) (N=**179583) | **P-values** |
|---|---|---|---|---|---|
| **Demographics** | | | | | |
| **Age, Mean (SD)** | 66.02(15.63) | 68.42(16.05) | 63.06(17.36) | 59.3(19.91) | < 0.0001 |
| **Gender (N, %)** | | | | | |
| Female | 20772 (40.82) | 8213 (39.34) | 26334 (48.13) | 91328 (50.86) | < 0.0001 |
| Male | 30110 (59.17) | 12660 (60.65) | 28371 (51.85) | 88232 (49.13) | |
| **Race (N, %)** | | | | | |
| White | 29091 (57.17) | 12771 (61.18) | 33843 (61.85) | 116777 (65.03) | < 0.0001 |
| Black | 13210 (25.96) | 5381 (25.78) | 9701 (17.73) | 30272 (16.86) | |
| Asian | 1375 (2.7) | 394 (1.89) | 1298 (2.37) | 4103 (2.28) | |
| Others | 895 (1.76) | 307 (1.47) | 1534 (2.8) | 4278 (2.38) | |
| No Information | 6318 (12.42) | 2022 (9.69) | 8338 (15.24) | 24153 (13.45) | |
| **Ethnicity (N, %)** | | | | | |
| Not Hispanic or Latino | 40282 (79.16) | 17180 (82.3) | 42204 (77.14) | 139564 (77.72) | < 0.0001 |
| Hispanic or Latino | 5555 (10.92) | 1640 (7.86) | 8559 (15.64) | 25466 (14.18) | |
| No Information | 5052 (9.93) | 2055 (9.84) | 3951 (7.22) | 14553 (8.1) | |
| **Co-morbid conditions (N, %)** | | | | | |
| History available | 37015 (72.74) | 16911 (81.01) | 38625 (70.59) | 139778 (77.83) | < 0.0001 |
| Among those with histories available | | | | | |
| *CVD* | 19602 (52.96) | 9527 (56.34) | 14162 (36.67) | 48639 (34.8) | < 0.0001 |
| DM | 5148 (43.1) | 16019 (43.28) | 11568 (29.95) | 35763 (25.59) | < 0.0001 |
| HF | 9500 (25.67) | 4655 (27.53) | 5864 (15.18) | 17658 (12.63) | < 0.0001 |
| HTN | 25321 (68.41) | 12125 (71.7) | 18152 (47.0) | 64755 (46.33) | < 0.0001 |
| **BMI, Mean (SD)** | 31.02(8.8) | 30.42(8.35) | 30.27(8.6) | 30.98(8.58) | |
| **Severity of illness (N, %)** | | | | | |
| Sepsis | 24169 (47.49) | 4560 (21.84) | 10631 (19.43) | 19146 (10.66) | <0.0001 |
| IMV | 18426 (36.21) | 1240 (5.94) | 8845 (16.17) | 3946 (2.2) | <0.0001 |
| Length of hospital stay, Mean, days (IQRs) | 17.17(18.91) | 7.54(8.04) | 14.41(15.29) | 6.46(7.05) | |

| | | | | | |
|---|---|---|---|---|---|
| **Medications (N, %)** | | | | | |
| VP | 17963 (35.3) | 1564 (7.49) | 10636 (19.44) | 9744 (5.43) | < 0.0001 |
| **Death (N, %)** | 18632 (36.61) | 3719 (17.82) | 11384 (20.81) | 12387 (6.9) | < 0.0001 |

**Abbreviations:**
DM (Diabetes Mellitus), HF (Heart Failure), HTN (Hypertension), CVD (Cardiovascular Disease), BMI (Body Mass Index, kg/m²), IMV (Invasive Mechanical Ventilation), VP (Vasopressor).

**Supplementary Table 3. Descriptive characteristics of each region**

| **Variables** | **Midwest** **N=**166456 | **South** **N=**56991 | **Northeast** **N=**56092 | **West** **N=**26522 | **P-values** |
|---|---|---|---|---|---|
| **Demographics** | | | | | |
| **Age, Mean (SD)** | 61.82(19.68) | 60.55(17.61) | 63.71(17.63) | 59.34(17.86) | < 0.0001 |
| **Gender (N, %)** | | | | | |
| Female | 80364 (48.28) | 27731 (48.66) | 26502 (47.25) | 12050 (45.43) | < 0.0001 |
| Male | 86081 (51.71) | 29250 (51.32) | 29586 (52.75) | 14456 (54.51) | |
| **Race (N, %)** | | | | | |
| White | 117236 (70.43) | 35965 (63.11) | 23787 (42.41) | 15494 (58.42) | < 0.0001 |
| Black | 31915 (19.17) | 14476 (25.4) | 9374 (16.71) | 2799 (10.55) | |
| Asian | 2981 (1.79) | 556 (0.98) | 2421 (4.32) | 1212 (4.57) | |
| Others | 4683 (2.81) | 602 (1.06) | 210 (0.37) | 1519 (5.73) | |
| No Information | 9641 (5.79) | 5392 (9.46) | 20300 (36.19) | 5498 (20.73) | |
| **Ethnicity (N, %)** | | | | | |
| Not Hispanic or Latino | 138783 (83.38) | 47068 (82.59) | 35018 (62.43) | 18361 (69.23) | < 0.0001 |
| Hispanic or Latino | 17048 (10.24) | 3271 (5.74) | 13660 (24.35) | 7241 (27.3) | |
| No Information | 10625 (6.38) | 6652 (11.67) | 7414 (13.22) | 920 (3.47) | |
| **Co-morbid conditions (N, %)** | | | | | |
| History available | 130364 (78.32) | 43909 (77.05) | 39315 (70.09) | 18741 (70.66) | < 0.0001 |
| Among those with histories available | | | | | |
| *CVD* | 47060 (36.1) | 20980 (47.78) | 16365 (41.63) | 7525 (40.15) | < 0.0001 |
| DM | 37321 (28.63) | 15732 (35.83) | 11650 (29.63) | 5791 (30.9) | < 0.0001 |
| HF | 20018 (15.36) | 8676 (19.76) | 6042 (15.37) | 2941 (15.69) | < 0.0001 |
| HTN | 63928 (49.04) | 26850 (61.15) | 19625 (49.92) | 9950 (53.09) | < 0.0001 |

| | | | | | |
|---|---|---|---|---|---|
| **Severity of illness (N, %)** | | | | | |
| Sepsis | 23900 (14.36) | 15781 (27.69) | 11419 (20.36) | 7406 (27.92) | <0.0001 |
| IMV | 12532 (7.53) | 8558 (15.02) | 6666 (11.88) | 4701 (17.72) | <0.0001 |
| **Medications (N, %)** | | | | | |
| VP | 15709 (9.44) | 8175 (14.34) | 9108 (16.24) | 6915 (26.07) | < 0.0001 |
| **AKI (N, %)** | 62889 (37.78) | 26816 (47.05) | 24148 (43.05) | 11992 (45.22) | p < 0.0001 |
| **Death (N, %)** | 22090 (13.27) | 10374 (18.2) | 9154 (16.32) | 4504 (16.98) | < 0.0001 |

**Abbreviations:**
DM (Diabetes Mellitus), HF (Heart Failure), HTN (Hypertension), CVD (Cardiovascular Disease), BMI (Body Mass Index, kg/m²), IMV (Invasive Mechanical Ventilation), VP (Vasopressor), AKI (Acute Kidney Injury).

**Supplementary Table 4. Descriptive characteristics of each time period**

| Variables | P1 N=46333 | P2 N=29110 | P3 N=98684 | P4 N=37419 | P5 N=37206 | P6 N=57309 | P-values |
|---|---|---|---|---|---|---|---|
| **Demographics** | | | | | | | |
| **Age, Mean (SD)** | 62.18(17.59) | 60.77(18.18) | 65.09(18.23) | 59.8(20.43) | 57.6(17.9) | 59.92(19.62) | < 0.0001 |
| **Gender (N, %)** | | | | | | | |
| Female | 21174 (45.7) | 14105 (48.45) | 46930 (47.56) | 18521 (49.5) | 18158 (48.8) | 27759 (48.44) | < 0.0001 |
| Male | 25150 (54.28) | 15002 (51.54) | 51743 (52.43) | 18894 (50.49) | 19044 (51.19) | 29540 (51.55) | |
| **Race (N, %)** | | | | | | | |
| White | 19765 (42.66) | 16869 (57.95) | 65754 (66.63) | 22374 (59.79) | 25707 (69.09) | 42013 (73.31) | < 0.0001 |
| Black | 11907 (25.7) | 6526 (22.42) | 15999 (16.21) | 8026 (21.45) | 7024 (18.88) | 9082 (15.85) | |
| Asian | 1683 (3.63) | 638 (2.19) | 2741 (2.78) | 1053 (2.81) | 474 (1.27) | 581 (1.01) | |
| Others | 1064 (2.3) | 1030 (3.54) | 2316 (2.35) | 733 (1.96) | 748 (2.01) | 1123 (1.96) | |
| No Information | 11914 (25.71) | 4047 (13.9) | 11874 (12.03) | 5233 (13.98) | 3253 (8.74) | 4510 (7.87) | |
| **Ethnicity (N, %)** | | | | | | | |
| Not Hispanic or Latino | 30303 (65.4) | 22184 (76.21) | 79305 (80.36) | 29072 (77.69) | 30924 (83.12) | 47442 (82.78) | < 0.0001 |
| Hispanic or Latino | 12052 (26.01) | 4770 (16.39) | 12656 (12.82) | 4869 (13.01) | 3065 (8.24) | 3808 (6.64) | |
| No Information | 3978 (8.59) | 2156 (7.41) | 6723 (6.81) | 3478 (9.29) | 3217 (8.65) | 6059 (10.57) | |
| **Co-morbid conditions (N, %)** | | | | | | | |
| History available | 30696 (66.25) | 21911 (75.27) | 76710 (77.73) | 28482 (76.12) | 28465 (76.51) | 46065 (80.38) | < 0.0001 |
| Among those with histories available | | | | | | | |
| *CVD* | 11340 (36.94) | 8370 (38.2) | 32992 (43.01) | 10792 (37.89) | 10064 (35.36) | 11340 (36.94) | < 0.0001 |
| DM | 9526 (31.03) | 6754 (30.82) | 24835 (32.38) | 8232 (28.9) | 7767 (27.29) | 13380 (29.05) | < 0.0001 |
| HF | 4651 (15.15) | 3389 (15.47) | 13709 (17.87) | 4321 (15.17) | 3977 (13.97) | 7630 (16.56) | < 0.0001 |
| HTN | 15393 (50.15) | 11127 (50.78) | 42535 (55.45) | 14276 (50.12) | 13496 (47.41) | 23526 (51.07) | < 0.0001 |
| **Severity of illness (N, %)** | | | | | | | |
| Sepsis | 12312 (26.57) | 5970 (20.51) | 17214 (17.44) | 6644 (17.76) | 7068 (19.0) | 9298 (16.22) | <0.0001 |
| IMV | 7550 (16.3) | 2894 (9.94) | 8404 (8.52) | 3426 (9.16) | 4156 (11.17) | 6027 (10.52) | < 0.0001 |
| **Medications (N, %)** | | | | | | | |
| VP | 8249 (17.8) | 3589 (12.33) | 10879 (11.02) | 4971 (13.28) | 5012 (13.47) | 7207 (12.58) | < 0.0001 |

| | | | | | | | |
|---|---|---|---|---|---|---|---|
| **AKI (N, %)** | 22505 (48.57) | 11700 (40.19) | 39656 (40.18) | 14058 (37.57) | 14675 (39.44) | 23251 (40.57) | <0.0001 |
| **Death (N, %)** | 9361 (20.2) | 4215 (14.48) | 16435 (16.65) | 4302 (11.5) | 4705 (12.65) | 7104 (12.4) | <0.0001 |

**Abbreviations:**
DM (Diabetes Mellitus), HF (Heart Failure), HTN (Hypertension), CVD (Cardiovascular Disease), BMI (Body Mass Index, kg/m²), IMV (Invasive Mechanical Ventilation), VP(Vasopressor ), AKI (Acute Kidney injury).

**Supplementary Table 5. Descriptive characteristics of each racial group.**

| Variables | White<br>N=192482 | Black<br>N=58564 | Asian<br>N=7170 | Others<br>N=7014 | P-values |
|---|---|---|---|---|---|
| **Demographics** | | | | | |
| **Age, Mean (SD)** | 64.04(19.01) | 57.66(17.74) | 61.65(17.65) | 55.12(17.87) | < 0.0001 |
| **Gender (N, %)** | | | | | |
| Female | 90029 (46.77) | 31773 (54.25) | 3244 (45.24) | 3084 (43.97) | < 0.0001 |
| Male | 102438 (53.22) | 26785 (45.74) | 3925 (54.74) | 3924 (55.95) | |
| **Ethnicity (N, %)** | | | | | |
| Not Hispanic or Latino | 164747 (85.59) | 53984 (92.18) | 6492 (90.54) | 2522 (35.96) | < 0.0001 |
| Hispanic or Latino | 13842 (7.19) | 874 (1.49) | 90 (1.26) | 3194 (45.54) | |
| No Information | 13893 (7.22) | 3706 (6.33) | 588 (8.2) | 1298 (18.51) | |
| **Co-morbid conditions (N, %)** | | | | | |
| History available | 152680 (79.32) | 45066 (76.95) | 4375 (61.02) | 4293 (61.21) | < 0.0001 |
| Among those with histories available | | | | | |
| *CVD* | 63951 (41.89) | 17485 (38.8) | 1466 (33.51) | 1132 (26.37) | < 0.0001 |
| DM | 43435 (28.45) | 16335 (36.25) | 1436 (32.82) | 1233 (28.72) | < 0.0001 |
| HF | 25547 (16.73) | 8414 (18.67) | 449 (10.26) | 416 (9.69) | < 0.0001 |
| HTN | 79339 (51.96) | 26164 (58.06) | 2145 (49.03) | 1578 (36.76) | < 0.0001 |
| **BMI, Mean (SD)** | 30.67(8.37) | 32.37(9.96) | 26.26(5.84) | 31.19(7.83) | < 0.0001 |
| **Severity of illness (N, %)** | | | | | |
| Sepsis | 35260 (18.32) | 11536 (19.7) | 1973 (27.52) | 1240 (17.68) | < 0.0001 |
| IMV | 18583 (9.65) | 6196 (10.58) | 1095 (15.27) | 725 (10.34) | <0.0001 |
| **Medications (N, %)** | | | | | |
| VP | 23504 (12.21) | 7327 (12.51) | 1288 (17.96) | 976 (13.92) | <0.0001 |
| **AKI (N, %)** | 75623 (39.29) | 28025 (47.85) | 3057 (42.64) | 2730 (38.92) | <0.0001 |
| **Death (N, %)** | 31218 (16.22) | 7256 (12.39) | 1125 (15.69) | 729 (10.39) | < 0.0001 |

**Abbreviations:**
DM (Diabetes Mellitus), HF (Heart Failure), HTN (Hypertension), CVD (Cardiovascular Disease), BMI (Body Mass Index, $kg/m^2$), IMV (Invasive Mechanical Ventilation), VP (Vasopressor), AKI (Acute Kidney Injury).

#### *Supplementary Figures*

**Supplementary Figure 1:** (a) Venn diagrams showing patients meeting different SCr-based AKI definitions **(b)** Comparison between the time of onset of AKI from the date of hospitalization and the length of hospitalization of patients (code-based AKI for all patients with AKI onset longer than the length of hospitalization). Color means the number of patients, darker the more patients. (**c)** Comparison of severity between patients meeting both AKI criteria *versus* those meeting only SCr criteria.

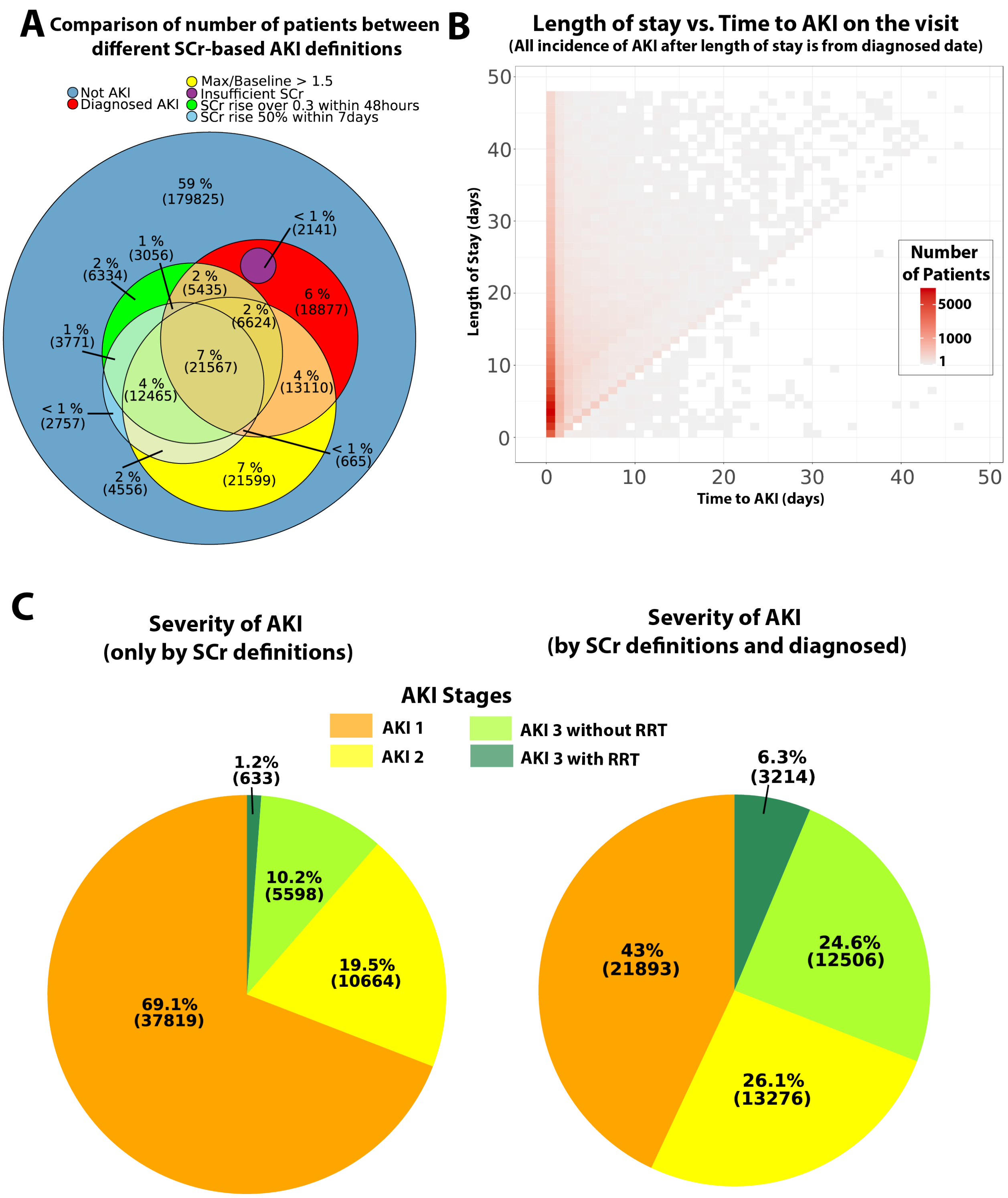

**Supplementary Figure 2: Survival of patients with and without AKI**

**(a)** First 60-day survival for patients diagnosed with AKI, but with insufficient SCr data to calculate a change. **(b)** Post-60-day survival for different AKI definitions. **(c)** First 60-day survival by KDIGO-based AKI stages and Code-based AKI. **(d)** Post-60-day survival by KDIGO-based AKI stages and Code-based AKI.

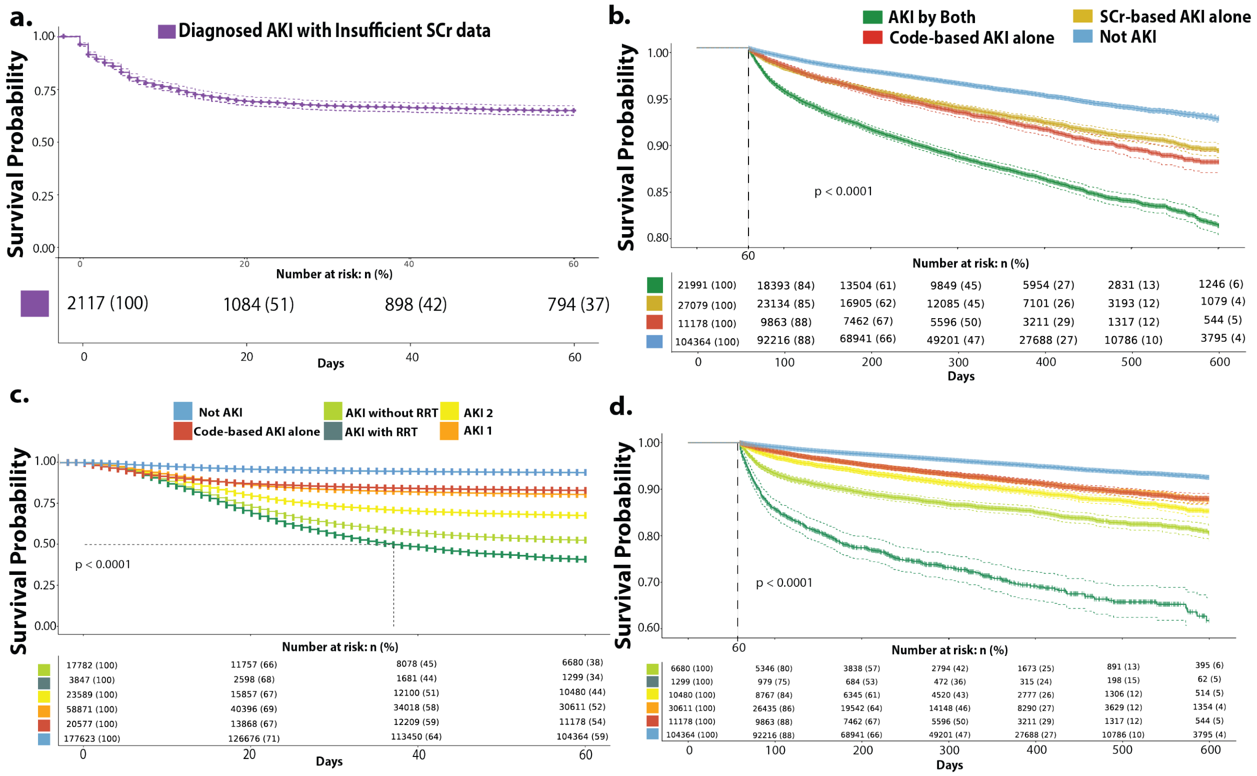

**Supplementary Figure 3: Mortality across time periods and regions**

**(a)** Comparison of univariate mortality rates (and 95% confidence intervals), between regional ad time frames. **(b)** Observed mortality rates (and 95% confidence intervals), within different severities of AKI, for six time periods.

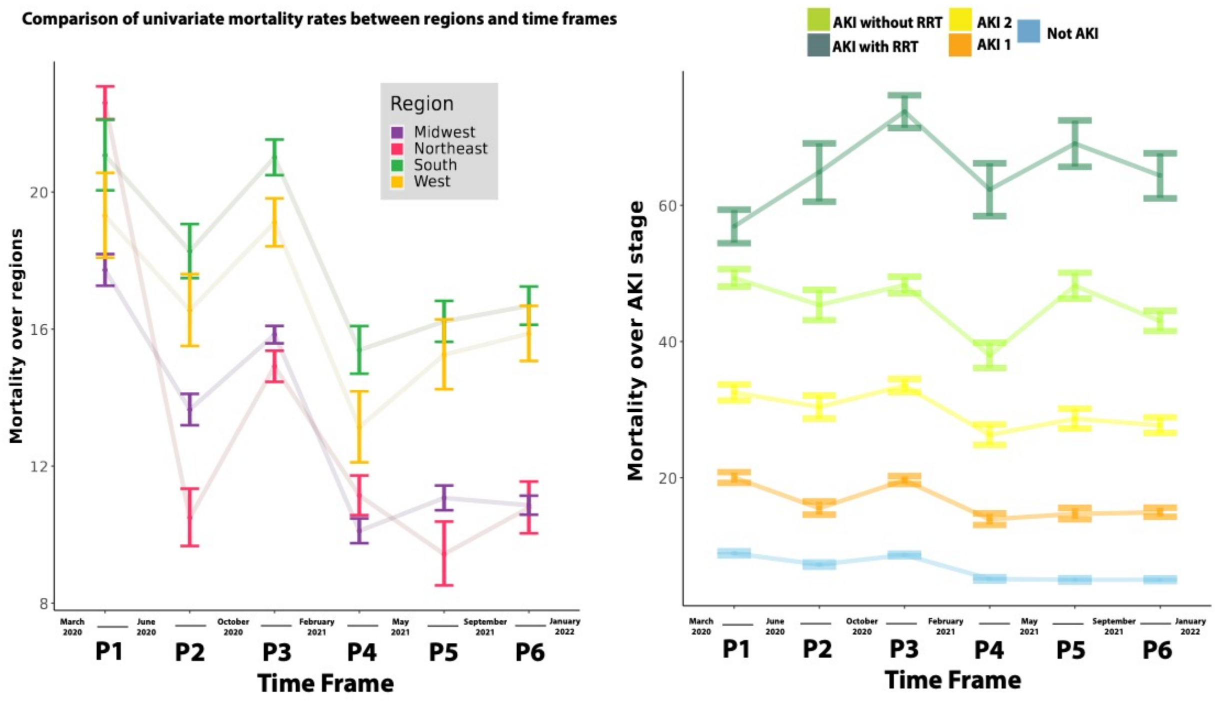

**Supplementary Figure 4: Multivariate analysis of COVID-19-related AKI risk in patients with BMI**

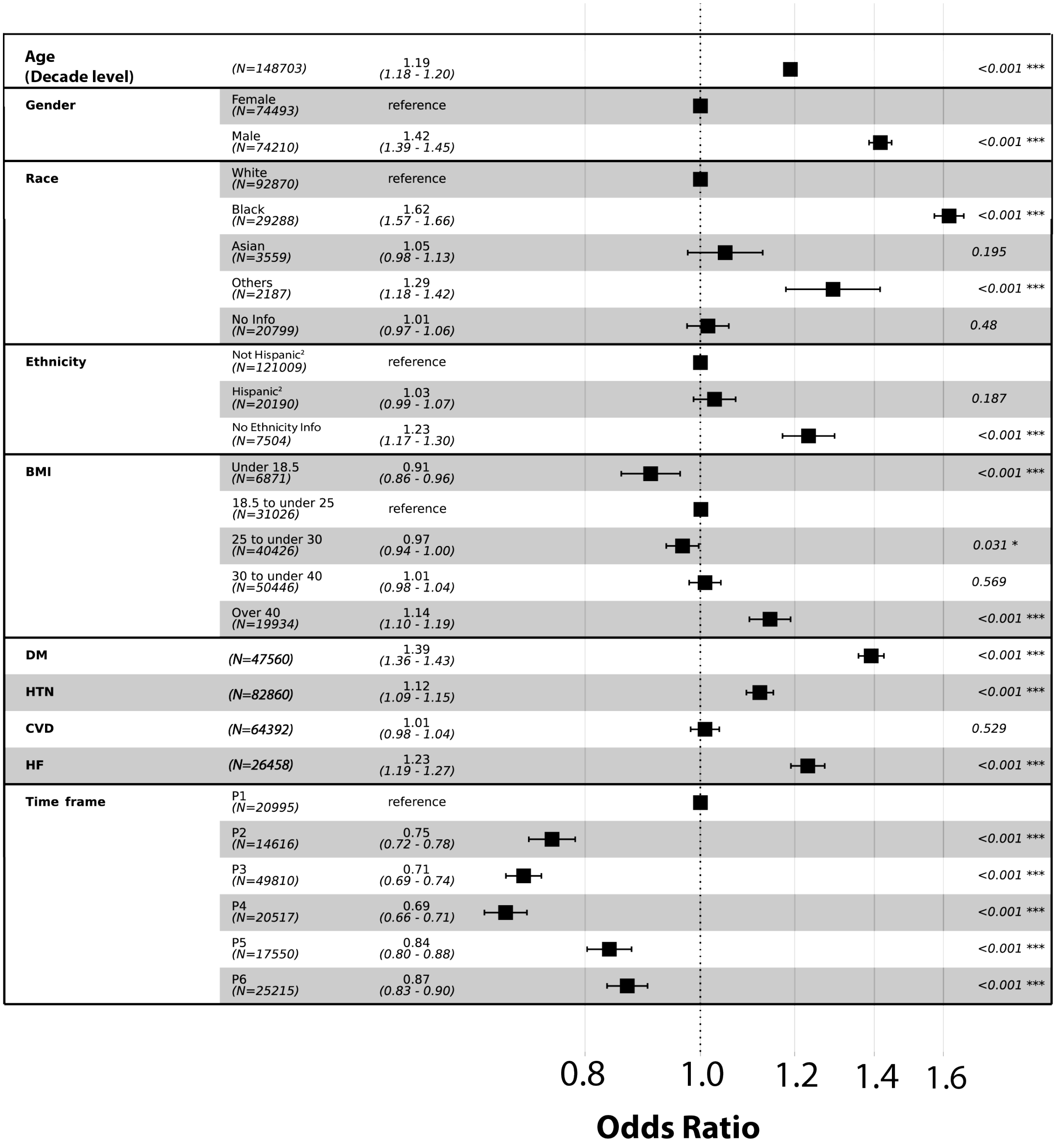

Age (Decade level)
(N=148703)
1.19 (1.18 - 1.20)
<0.001 ***
Gender
Female (N=74493)
reference
Male (N=74210)
1.42 (1.39 - 1.45)
<0.001 ***
Race
White (N=92870)
reference
Black (N=29288)
1.62 (1.57 - 1.66)
<0.001 ***
Asian (N=3559)
1.05 (0.98 - 1.13)
0.195
Others (N=2187)
1.29 (1.18 - 1.42)
<0.001 ***
No Info (N=20799)
1.01 (0.97 - 1.06)
0.48
Ethnicity
Not Hispanic[2] (N=121009)
reference
Hispanic[2] (N=20190)
1.03 (0.99 - 1.07)
0.187
No Ethnicity Info (N=7504)
1.23 (1.17 - 1.30)
<0.001 ***
BMI
Under 18.5 (N=6871)
0.91 (0.86 - 0.96)
<0.001 ***
18.5 to under 25 (N=31026)
reference
25 to under 30 (N=40426)
0.97 (0.94 - 1.00)
0.031 *
30 to under 40 (N=50446)
1.01 (0.98 - 1.04)
0.569
Over 40 (N=19934)
1.14 (1.10 - 1.19)
<0.001 ***
DM
(N=47560)
1.39 (1.36 - 1.43)
<0.001 ***
HTN
(N=82860)
1.12 (1.09 - 1.15)
<0.001 ***
CVD
(N=64392)
1.01 (0.98 - 1.04)
0.529
HF
(N=26458)
1.23 (1.19 - 1.27)
<0.001 ***
Time frame
P1 (N=20995)
reference
P2 (N=14616)
0.75 (0.72 - 0.78)
<0.001 ***
P3 (N=49810)
0.71 (0.69 - 0.74)
<0.001 ***
P4 (N=20517)
0.69 (0.66 - 0.71)
<0.001 ***
P5 (N=17550)
0.84 (0.80 - 0.88)
<0.001 ***
P6 (N=25215)
0.87 (0.83 - 0.90)
<0.001 ***
0.8
1.0
1.2
1.4
1.6
Odds Ratio

**Supplementary Figure 5: Multivariable analysis of mortality in patients with BMI**

**(a)** Multivariable survival analysis of 113,216 patients using the Cox Proportional Hazards (CoxPH) model BMI and comorbidity. **(b)** CoxPH multivariate analysis including only HTN with BMI.

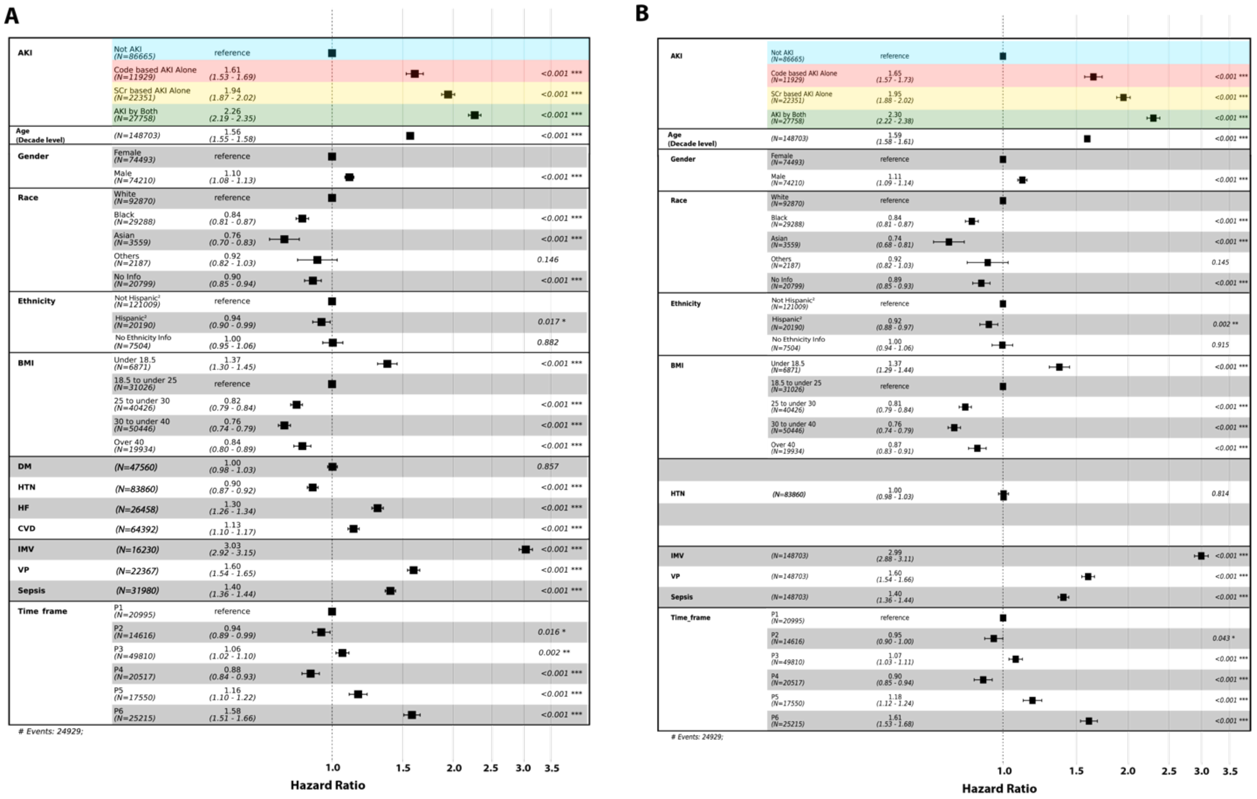

### Supplementary Figure 6: Follow-up comparison between Race groups and Venn-diagram of Comorbidities (HTN, DM, HF, CVD).

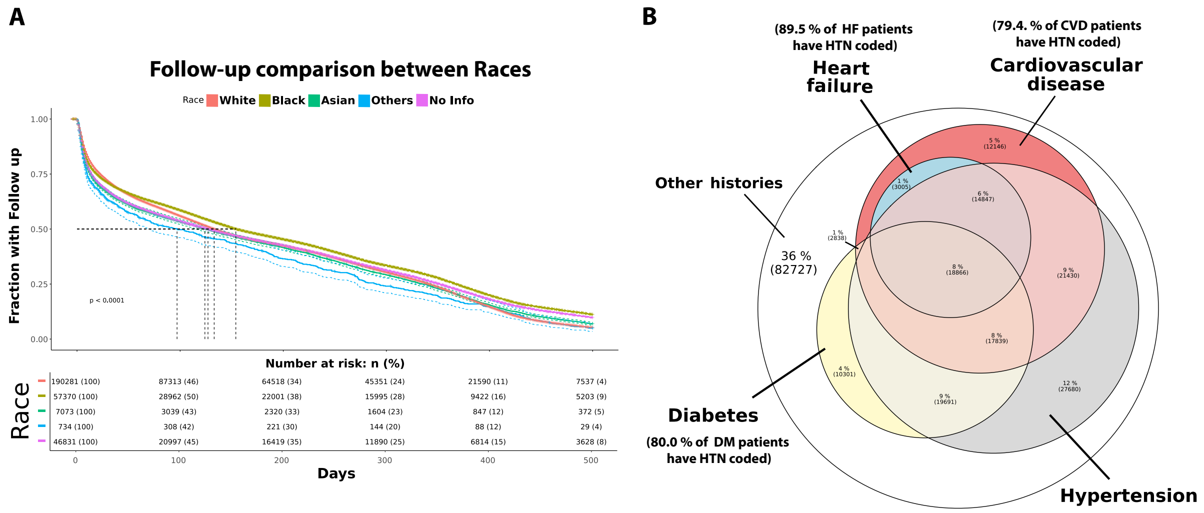
